# The individual and global impact of copy number variants on complex human traits

**DOI:** 10.1101/2021.08.10.21261839

**Authors:** Chiara Auwerx, Maarja Lepamets, Marie C. Sadler, Marion Patxot, Miloš Stojanov, David Baud, Reedik Mägi, Eleonora Porcu, Alexandre Reymond, Zoltán Kutalik

## Abstract

Copy number variations (CNVs) have been involved in multiple genomic disorders but their impact on complex traits remains understudied. We called CNVs in the UK Biobank and performed genome-wide association scans (GWASs) between the copy-number of CNV-proxy probes and 57 continuous traits, revealing 131 signals spanning 47 phenotypes. Our analysis recapitulated well-known associations (1q21 and height), revealed the pleiotropy of recurrent CNVs (26 traits for 16p11.2-BP4-BP5), and suggested new gene functionalities (*MARF1* in female reproduction). Forty CNV signals overlapped known GWAS loci (*RHD* deletion and hematological traits). Conversely, others overlapped Mendelian disorder regions, suggesting variable expressivity and a broad impact of these loci, as illustrated by signals mapping to Rotor syndrome (*SLCO1B1/3*), renal cysts and diabetes (*HNF1B*), or Charcot-Marie-Tooth (*PMP22*) loci. The total CNV burden negatively impacted 35 traits, leading to increased adiposity, liver/kidney damage, and decreased intelligence and physical capacity. Thirty traits remained burden-associated after correcting for CNV-GWAS signals, pointing to a polygenic CNV-architecture. The burden negatively correlated with socio-economic indicators, parental lifespan, and age (survivorship proxy), suggesting that CNVs contribute to decreased longevity. Together, our results showcase how studying CNVs can reveal new biological insights, emphasizing the critical role of this mutational class in shaping complex traits.

## INTRODUCTION

With the advent of genome-wide associations studies (GWASs), the polygenic architecture of complex human traits has become apparent (Canela-Xandri et al., 2018; Visscher et al., 2017; Watanabe et al., 2019). Still, single nucleotide polymorphisms (SNPs) do not explain the totality of observed phenotypic variability – a phenomenon referred to as *missing heritability* – and one proposed explanation is the contribution of additional types of genetic variants, such as copy number variants (CNVs) (Manolio et al., 2009).

Characterized by the deletion or duplication of DNA fragments ≥ 50 bp (Sudmant et al., 2015), CNVs represent a highly diverse mutational class that due to their possibly large size constitute potent phenotypic modifiers that act through e.g. gene dosage sensitivity, truncation or fusion of genes, unmasking of recessive alleles, or disruption of *cis*-regulatory elements (Shaikh, 2017). Hence, CNVs have been acknowledged to play an important role in human diseases and were identified as the genetic etiology of 65 rare and debilitating genomic syndromes by DECIPHER (Firth et al., 2009; see Web Resources). However, early GWASs failed to establish clear links between CNVs and complex traits and diseases (Craddock et al., 2010; Kathiresan et al., 2009). Several factors, specific to CNV-GWAS, contributed to these negative results, such as the low frequency and variable breakpoints of CNVs in the population, as well as uncertainty and low resolution of CNV calls originating from genotyping microarrays (Valsesia et al., 2013). In recent years, methodological development, as well as the creation of large biobanks has allowed to bypass some of these hurdles. Focusing on a curated set of CNVs, a series of studies characterized the impact of well-established pathogenic CNVs on cognitive performance (Kendall et al., 2017), physical measurements (Owen et al., 2018; Warland et al., 2019), common medical conditions (Crawford et al., 2019; Kendall et al., 2019), and blood biomarkers (Bracher-Smith et al., 2019). Alternatively, unbiased genome-wide (GW) studies have been conducted (Aguirre et al., 2019; Li et al., 2020; Macé et al., 2017; Sinnott-Armstrong et al., 2021), involving loci not covered by targeted approaches and adding to the growing body of evidence implicating CNVs in complex traits. Notably, a recent study made use of the UK Biobank (UKBB) (Bycroft et al., 2018) to assess the impact of CNVs on over 3’000 phenotypes, providing the research community with the largest population-based CNV-to-phenotype resource to date (Aguirre et al., 2019). Using an independent CNV calling and association pipeline and focusing on a set of 57 medically relevant continuous traits, we here confirm previously established associations, reveal new biological insight through in-depth analysis of particular CNV-trait pairs, and expose a nuanced role of CNVs along the rare versus common disease spectrum, suggesting that the deleterious impact of CNVs contributes to decreased longevity in the general population.

## RESULTS

### The CNV landscape of the UK Biobank

PennCNV (Wang et al., 2007) was used to call autosomal and chrX CNVs in 332’935 unrelated white British UKBB participants with no reported blood malignancy. Calls were processed by a pipeline that excluded 1’413 CNV outlier samples and attributed a probabilistic quality score (QS) to each CNV (Macé et al., 2016). Out of 1’329’290 identified CNVs, 176’870 high confidence CNVs with |QS| ≥ 0.5 were retained for follow-up analyses (Figure S1A). Overall, 129’263 (39%) participants carried at least one high confidence CNV and 34’804 (10%) carried more than one (Figure S1B). In samples with ≥ 1 CNV, the total length of affected bases ranged between 217 bp and 14.2 Mb, with a median of 292 kb (Figure S1C). Analyzing the global CNV burden of the cohort, 70% was caused by duplications, which were both more numerous (54%) and 213 kb longer, on average, than deletions (Figure S1D). No differences in CNV burden, measured as the number of Mb or genes affected by CNVs, was detected across sexes (two-sided, unpaired Wilcoxon rank sum test: *p*_*Mb*_ = 0.793; *p*_*Genes*_ = 0.748). This contrasts with the excess of deleterious CNVs reported in females with neuro-psychiatric/developmental disorders (Gilman et al., 2011; Jacquemont et al., 2014; Levy et al., 2011; Sanders et al., 2015), suggesting that this observation is trait-dependent.

To bypass issues related to inter-individual variability in recurrent CNV break points, CNV calls were transformed to the probe level for frequency calculation (Table S1) (Macé et al., 2017). A large fraction of the genome was subjected to CNVs as 662’247 probes (82%) were found in a copy number-altered state in at least one participant, even if 81% of these had a CNV frequency ≤ 0.005% (n ≤ 16). The fraction of never-deleted probes (43%) was 1.7x higher than the fraction of never-duplicated probes (26%) and with some notable exceptions, deletion frequencies tended to be lower than duplication frequencies (Figure 1). For most loci with high CNV frequency, duplication and deletion frequencies did not mirror each other (Figure 1). Overall, these results are in line with the common paradigm that CNVs are individually rare but collectively common (Abel et al., 2020; Aguirre et al., 2019; Collins et al., 2020).

**Figure 1.**
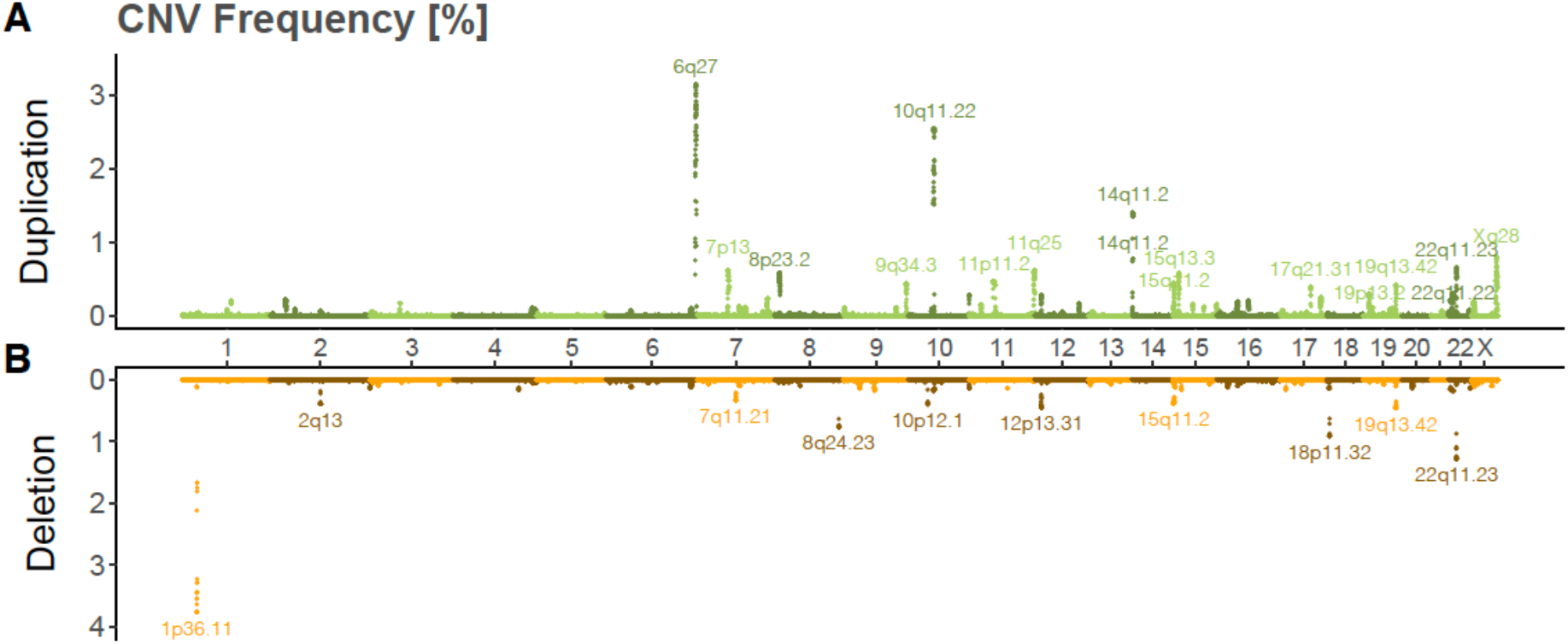
The CNV frequency landscape in the UKBB. Miami plot of high confidence probe-level **(A)** duplication and **(B)** deletion frequencies [%] in the UKBB. Loci with duplication frequency ≥ 0.3% or deletion frequency ≥ 0.2% are labeled with cytogenic bands.

### The pleiotropic impact of recurrent CNVs

To assess the phenotypic impact of the UKBB CNV landscape, we selected 57 medically relevant phenotypes – including anthropometric traits, cardio-pulmonary assessments, hematological measurements, blood biomarkers, neuronal functions, and sex-specific attributes – with presumed high heritability (Table S3; Figure S2). GWASs were performed between the copy number (CN) of pruned (r^2^ > 0.9999) CNV-proxy probes with a CNV frequency ≥ 0.005% and aforementioned traits according to a mirror (28’257 probes; Figure 2A) and two CNV-type specific models: a duplication-only (14’070 probes; Figure 2B) and deletion-only (9’936 probes; Figure 2C) association model. Summary statistics are available as Supplementary data. Stepwise conditional analysis narrowed signals down to 88, 51, and 71 GW-significant associations (*p* ≤ 0.05/11’804 = 4.2×10^−6^; see Methods) for the mirror, duplication-only, and deletion-only models, respectively. These were combined into 131 independent signals spanning 47 phenotypes (Figure 2D; Table S4). Among signals identified through the mirror model, 63 (73%) replicated with either type-specific model, often reflecting the most common CNV type (Figure 2D top) and 5 (6%) replicated with both type-specific models, providing examples of “true mirror” effects (*i*.*e*. opposite impact of duplications and deletions), as exemplified by the association between the CN of a pseudoautosomal (Xp22.33) CNV region (CNVR) encompassing *SHOX* (short-stature homeobox) and increased height (MIM: 127300, 249700). Alternatively, partially overlapping signals between decreased forced vital capacity or grip strength and the DiGeorge/velocardiofacial syndrome locus (MIM: 188400, 192430) 22q11.21 low copy repeat (LCR) A-B (deletion-only) and 22q11.21 LCR A-D (mirror and duplication-only) hinted at U-shaped effects (*i*.*e*. deletion and duplication shift the phenotype in the same direction), demonstrating the existence of different mechanisms of gene dosage for at least one of the encompassed genes.

**Figure 2.**
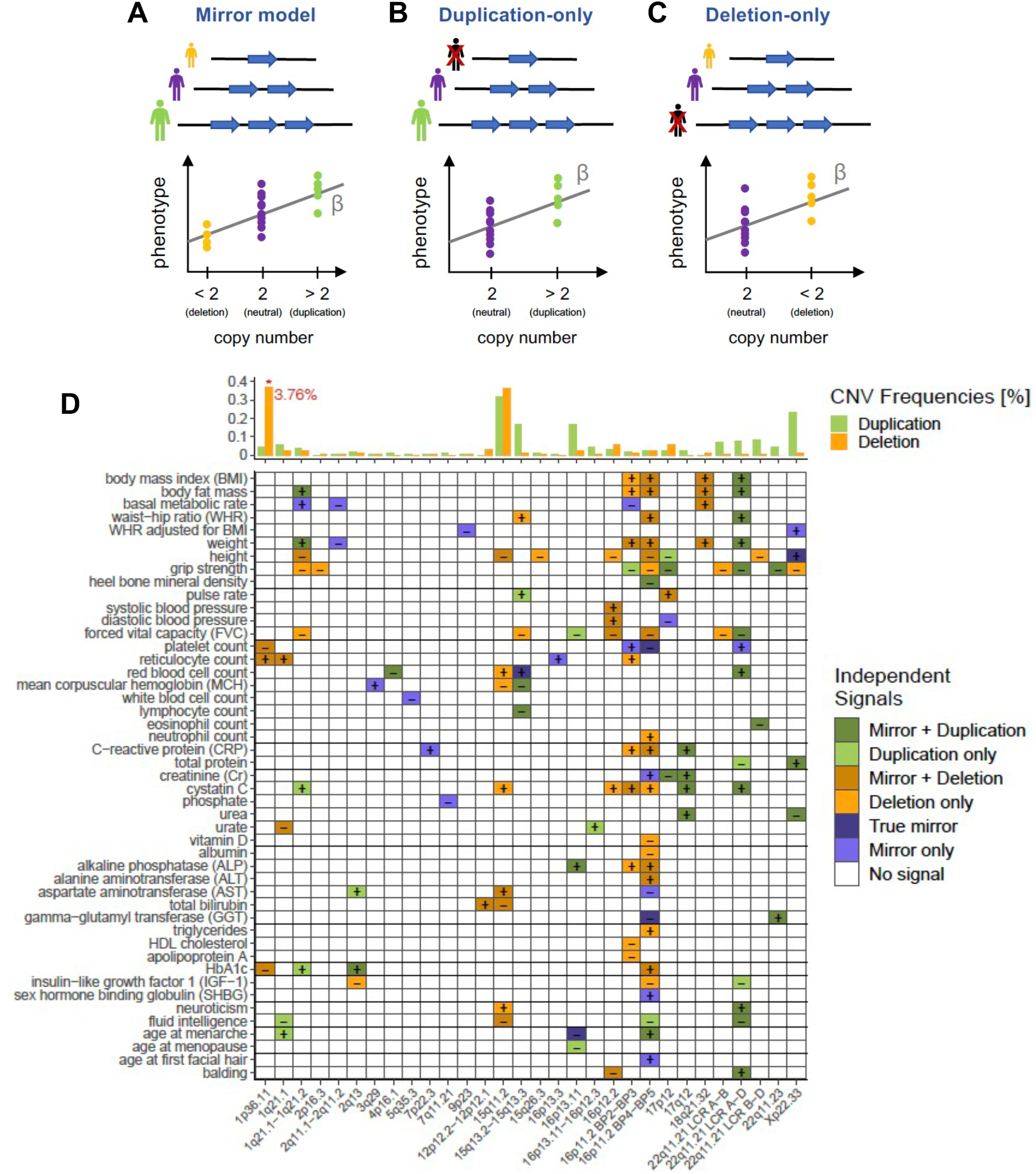
CNV-GWAS roadmap of the UKBB. CNV-GWAS association models: **(A)** The mirror model assumes an equal-sized but opposite-direction effect of deletion and duplication and estimates the impact of each additional copy number; **(B)** The duplication-only model disregards deletion carriers and estimates the effect of a duplication; **(C)** The deletion-only model disregards duplication carriers and estimates the effect of a deletion. **(D)** Independent GW-significant associations between CNVRs (x-axis; as cytogenic bands) and phenotypes (y-axis). Colors represent the model(s) through which the association was detected – dark green: mirror and duplication-only; light green: duplication-only; dark orange: mirror and deletion-only; light orange: deletion-only; dark purple: mirror, duplication-only, and deletion-only; light purple: mirror; white: none - and signs show directionality, so that the duplication (greens), deletion (oranges), or CN (purples) of a CNVR correlated with an increase (“+”) or decrease (“-”) in the associated trait. 16p11.2 and 22q11.21 (LCRB at chr22: 20’400’000) were split to distinguish CNVRs. For each CNVR, average duplication (green) and deletion (orange) frequencies [%] of the lead probe according to the most significant model are depicted at the top. Deletion frequency of 1p36.11 was truncated from 3.76%.

Most signals involved large (mean = 817 kb) recurrent CNVRs and we confirm multiple well-established associations, such as the negative impact of the 1q21.1-1q21.2 deletion on height (Bernier et al., 2016; Brunetti-Pierri et al., 2008; Mefford et al., 2008), the negative correlation between body mass index (BMI) and the CN of 16p11.2 BP4-BP5 (Bochukova et al., 2010; Conrad et al., 2010; Jacquemont et al., 2011) and 16p11.2 BP2-BP3 (Bachmann-Gagescu et al., 2010; Bochukova et al., 2010; Loviglio et al., 2017), or the more recently reported positive association between 16p11.2 BP4-BP5’s CN and age at menarche (Männik et al., 2019). In addition, our results revealed the broad pleiotropic impact of these loci. While the median CNVR affected 3 traits, other loci, such as 16p11.2 BP4-BP5, 22q11.21, or 16p11.2 BP2-BP3, associated with 26, 16, and 12 phenotypes, respectively, emphasizing the potent role of CNVs as phenotypic modifiers.

### Replication in the Estonian Biobank

We sought to replicate identified signals in an independent cohort, the Estonian Biobank (EstBB) (Leitsalu et al., 2015). CNV data were available for 89’516 unrelated individuals (Table S2) but phenotypic measurements, originating from national health registries, were only available for a limited subset of participants, ranging between ∼60’000 for anthropometric measurements, to < 1’000 for specialized biomarkers (Table S3). Restricting ourselves to autosomal signals with sample size ≥ 2’000 and ≥ 1 CNV carrier, data were available for 61 (47%) CNVR-trait pairs (Table S4; Figure 3A). Six signals replicated with Bonferroni correction (5%) for multiple testing (*p* ≤ 0.05/61 = 8.2×10^−4^; Figure 3B) and we observed 7.2x more nominally significant signals than expected by chance (22 signals; binomial test: *p* = 7.8×10^−14^; Figure S3A), with effect size estimates following closely the ones detected in the UKBB (Figure 3). Given the low sample sizes, simulations were conducted to assess the power of the replication study. Assuming effect sizes matching those observed in the UKBB and using CNV frequencies and sample sizes reflective of the EstBB, the average replication power was estimated at 5.5% (at α = 0.05/61; Figure S3B). This corresponds to an expected number of replicated signals of 3.4, slightly below the six observed, and argues in favor of the robustness of the original CNV-GWAS findings.

**Figure 3.**
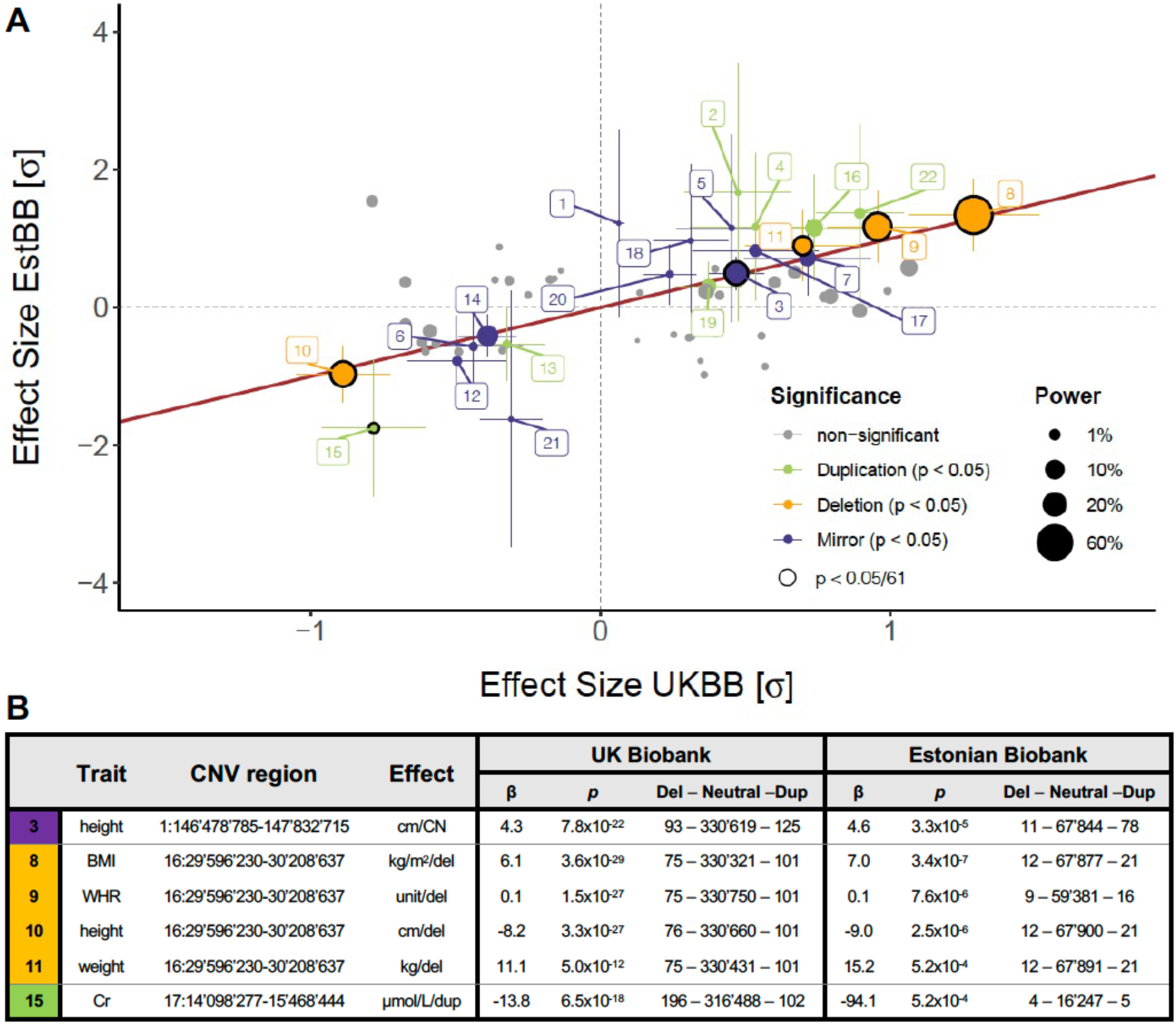
Replication of CNV-GWAS signals in the EstBB. **(A)** EstBB (y-axis) versus UKBB (x-axis) standardized effect sizes. The identity line is shown in red; Size reflects power at α = 0.05/61; Non-significant signals (*p* > 0.05) are in grey; Nominally significant signals (*p* ≤ 0.05) with 95% confidence intervals are colored according to replication models: mirror (purple), duplication-only (green), or deletion-only (orange); Multiple-testing correction surviving signals (*p* ≤ 8.2 × 10^−4^) are circled in black and listed in **(B)** with the first column’s color corresponding to the association model and numbers matching labels in **(A)**, effect sizes (β; unit in the effect column) and *p*-values (*p*) for the UKBB and EstBB GWAS, along with the number of individuals with available phenotypic data carrying a deletion, no CNV, or a duplication overlapping the CNVR. Labels indicate: 1p36.11 – (1) platelet count; 1q21.1-1q21.2 – (2) HbA1c, (3) height; 1q21.1 – (4) age at menarche; 16p11.2 BP2-BP3 – (5) platelet count, (6) weight; 16p11.2 BP4-BP5 – (7) age at menarche, (8) BMI, (9) WHR, (10) height, (11) weight, (12) ALT; 16p13.11 – age at (13) menopause and (14) menarche; 17p12 – (15) SCr; 17q12 – (16) SCr, (17) CRP; 22q11.21 LCR A-D – (18) platelet count, (19) BMI, (20) weight; 22q11.21 LCR B-D – (21) eosinophil count; 22q11.23 – (22) GGT.

### CNVs as modifiers of complex traits

To assess whether CNV-GWAS signals mapped to loci previously identified through SNP-GWAS for the same trait, we annotated CNVRs with associations reported by the NHGRI-EBI GWAS catalog (Buniello et al., 2019). Forty CNV signals (31%) overlapped with a corresponding SNP signal (Table S4), corroborating the role of these regions in the genetic architecture of analyzed traits.

A first example is the association between the 382 kb 1q21.1 deletion (chr1:145’383’239-145’765’206) and decreased serum urate levels (βdel = −48.32 µmol/L; *p* = 5.8×10^−13^; Figure 4A). The rearranged interval encompasses 15 genes (Figure S4), including *PDZK1*, which encodes a urate transporter scaffold protein (Anzai et al., 2004) and was associated with serum urate in numerous SNP-GWAS (Kolz et al., 2009; Köttgen et al., 2013; Sulem et al., 2011; Yang et al., 2010). Recently, *in vitro* experiments elucidated the mechanism through which the urate-increasing T allele of rs1967017 led to increased *PDZK1* expression (Ketharnathan et al., 2018), while the *PDZK1* protein truncating variant rs191362962 was found to associate with decreased serum urate (Sinnott-Armstrong et al., 2021), both suggesting that decreased *PDZK1* expression – an expected outcome of *PDZK1* deletion – decreases serum urate levels. Dividing deletion carriers into groups harboring either a full (start < 145.6 Mb) or a partial (start ≥ 145.6 Mb) deletion revealed that the small deletion, which encompasses *PDZK1* and three other genes (Figure S4), was sufficient to alter serum urate levels (two-sided t-test: *p* = 0.92; Figure 4A).

**Figure 4.**
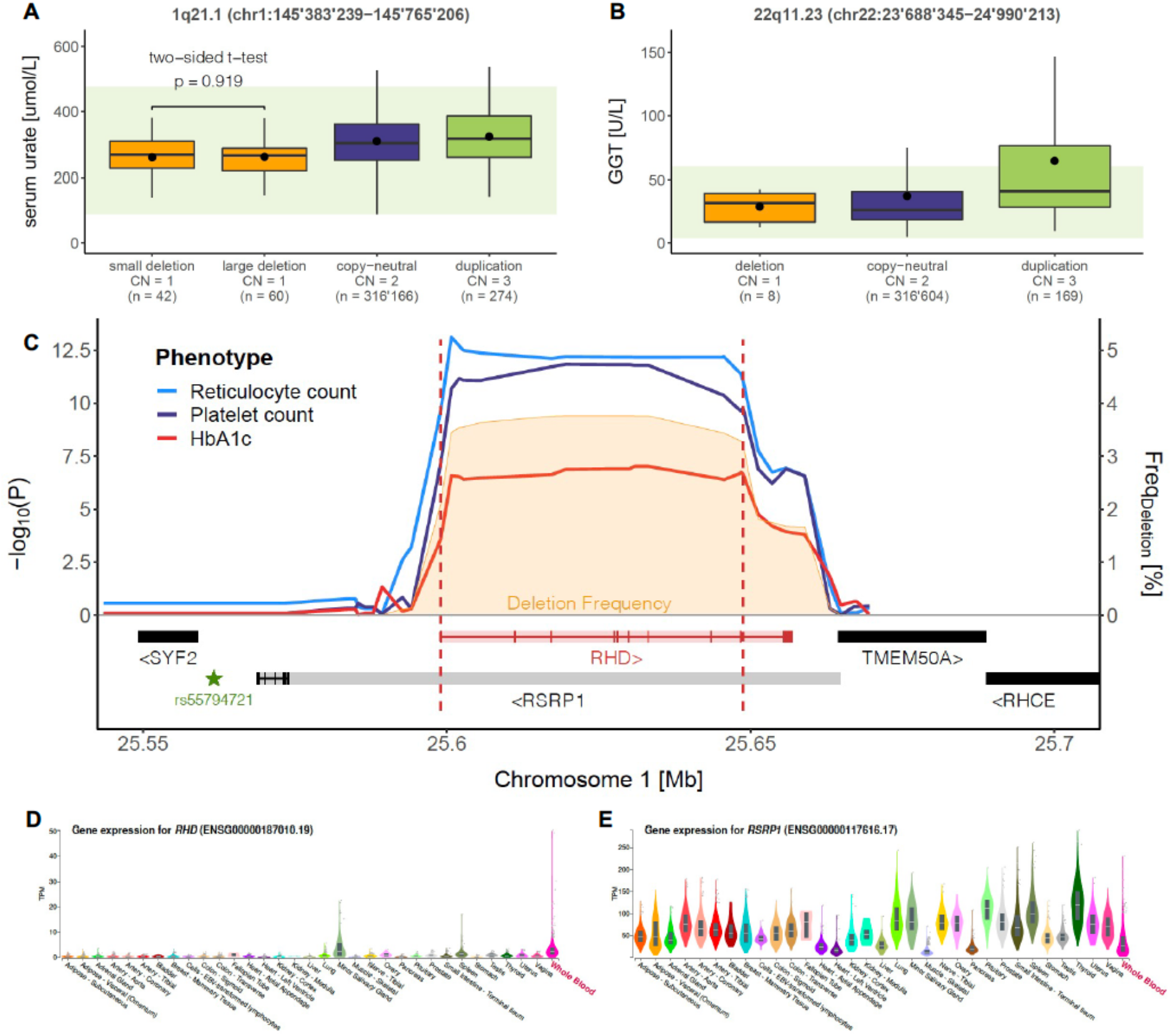
CNV-GWAS associations at SNP-GWAS loci. Boxplots representing levels of **(A)** serum urate in individuals with 1q21.1 small or large deletion, copy-neutrality, or duplication, and **(B)** GGT in individuals with 22q11.23 deletion, copy-neutrality, or duplication. CNVR coordinates are at the top; Copy number (CN) and sample size are reported for each category; Dots show the mean; Outliers are not shown; Green bands show normal clinical range for **(A)** serum urate: 89-476 μmol/L and **(B)** GGT: 4-6 U/L. **(C)** Association plot for the 1p36.11 deletion. Red dashed lines delimit the deletion-only CNVR (chr1: 25’599’041-25’648’747); Left y-axis shows the negative logarithm of association *p*-value for reticulocyte count (blue), platelet count (purple), and HbA1c (red); Right y-axis shows deletion frequency [%] (orange); Encompassed genes are at the bottom; Retained exons for the most strongly expressed isoform in whole blood are shown for *RHD* (ENST00000328664) and *RSRP1* (ENST00000243189), with shaded color representing the full gene sequence; Star indicates the *RHD* and *RSRP1* eQTL rs55794721. GTEx v8 gene expression in 33 tissues for **(D)** *RHD* and **(E)** *RSRP1*. Brain, cervix, esophagus, and skin are not shown for visibility. Whole blood is shown with a red label.

A second example is the association between the 1.3 Mb-long 22q11.23 duplication (chr22:23’688’345-24’990’213) and increased *γ*-glutamyl transferase (GGT; β_dup_ = 37.2 U/L; *p* = 9.3×10^−32^; Figure 4B). The region harbors several independent SNP-GWAS signals for GGT (Chambers et al., 2011; Gurdasani et al., 2019; Kamatani et al., 2010; Pazoki et al., 2021; Seo et al., 2020; Sinnott-Armstrong et al., 2021; Yuan et al., 2008), as well as 4 genes involved in glutathione metabolism (KEGG pathway hsa00480), including *GGT1* and *GGT5* themselves (Figure S5A), suggesting that an additional copy of these genes associates with increased levels of the encoded protein. As multiple factors can elevate GGT levels (Dufour et al., 2000), binomial tests were used to verify that the 180 duplications carriers were not enriched for GGT-altering drug usage (*p* = 0.55), high alcohol consumption (*p* = 0.85), heart failure (*p* = 0.23), or cancer (*p* = 1) and other diseases (*p* = 0.64) of the liver, gallbladder, and bile ducts, as compared to control individuals. Furthermore, visualization of GGT levels in individuals with two or three copies of the CNVR showed that the 22q11.23 duplication increased serum GGT independently of and additively to other GGT-increasing factors (Figure S5B-F).

Third, we focused on the most frequent CNV in our cohort, the 50 kb 1p36.11 deletion (chr1:25’599’041-25’648’747, frequency = 3.76%; Figure 1A), which encompass *RHD* and *RSRP1* and associated with increased reticulocyte count (β_del_ = 2.7×10^9^ cells/L; *p* = 7.8×10^−14^), decreased platelet count (β_del_ = −3.7×10^9^ cells/L; *p* = 1.4×10^−12^), and decreased glycated hemoglobin (HbA1c; β_del_ = −0.3 mmol/mol; *p* = 9.3×10^−8^) (Figure 4C). Overlap with SNP-GWAS signals for various hematological traits (Astle et al., 2016; Vuckovic et al., 2020) prompted the investigation of the expression of these genes in whole blood (WB). Tissue-specific transcriptomic data from the GTEx project v8 (Aguet et al., 2020; see Web Resources) revealed that *RHD* (Rhesus (Rh) blood group D antigen), a protein whose presence/absence on erythrocyte cell membranes is critical in determining an individual’s Rh blood group (Avent and Reid, 2000), was almost exclusively expressed in WB (Figure 4D), whereas *RSRP1* was ubiquitously expressed, with low expression in WB (Figure 4E). Selecting *RHD*’s (ENST00000328664) and *RSRP1*’s (ENST00000243189; Figure S6A) most highly expressed isoforms in WB, we mapped exons to the association plot, showing that ENST00000243189 (*RSRP1*) does not overlap the CNVR, in contrast to ENST00000328664 (*RHD*), which is fully encompassed by it (Figure 4C). To strengthen the hypothesis that *RHD-*CN is causal for the observed associations, we used transcriptome-wide Mendelian randomization (TWMR; Table S5) to establish a directionally concordant causal link between *RHD* expression and reticulocyte count (α_TWMR_ = −0.013 *p* = 1.6×10^−4^; Figure S6B), platelet count (α_TWMR_ = 0.031, *p* = 2.3×10^−9^; Figure S6C), and HbA1c (α_TWMR_ = 0.017, *p* = 3.5×10^−7^; Figure S6D). The same analysis was performed for *RSRP1* and while results were directionally concordant and significant, the gene had insufficient number of instruments (3) for robust TWMR (Figure S6E-G). Note that the signals for both *RHD* and *RSRP1* were driven by a strong upstream expression quantitative locus (eQTL) (rs55794721) (Figure S6B-G). To gauge the generalizability of these results, we tested whether similar trends could be observed in individuals with an Rh^-^ blood type. As this information was not available for the UKBB, we turned to a maternity cohort from the Lausanne University Hospital (CHUV). Despite low samples sizes, we observed concordant trends of increased reticulocyte count (β_*Rh-*_ = 1.07 °/oo; *p*_*one-sided*_ = 0.134; n = 741) and decreased platelet count (β_*Rh-*_ = −2.8×10^9^ cells/L; *p*_*one-sided*_ = 0.126; n = 5’034) and HbA1c levels (β_*Rh-*_ = −0.22%; *p*_*one-sided*_ = 0.050; n = 418) in Rh^-^ women (Table S6). Overall, these examples illustrate how studying CNVs at SNP-GWAS loci can shed light on causal genes and shared genetic mechanisms.

### CNVs at Mendelian disorder loci

Despite the lower-than-average disease burden of UKBB participants (Fry et al., 2017), we identified several associations comprising loci involved in Mendelian disorders. For instance, the heterozygous 395 kb 12p12.2-p12.1 deletion (chr12:21’008’080-21’403’457), which is associated with a non-pathological increase in total bilirubin (β_del_ = 3.1 µmol/L, *p =* 2.2×10^−13^; Figure 5A) and contains SNP-GWAS signals for bilirubin levels (Bielinski et al., 2011; Dai et al., 2013; Johnson et al., 2009; Kang et al., 2010; Sanna et al., 2009; Sinnott-Armstrong et al., 2021), overlaps the locus causing Rotor syndrome (MIM: 237450), an extremely rare disorder whose main clinical manifestation is hyperbilirubinemia. Rotor syndrome is caused by the homozygous disruption of *SLCO1B1* and *SLCO1B3* (Figure S7) (Van De Steeg et al., 2012), which encode for the hepatic transporters OATP1B1 and OATP1B3, respectively, involved in the uptake of various drugs and metabolic compounds, including bilirubin (Smith et al., 2005). Concordantly, UKBB participants diagnosed with Rotor syndrome or the related and more common Dubin-Johnson syndrome (MIM: 237500) present with above-normal levels of total bilirubin (Figure 5A). Interestingly, individuals carrying a partial deletion that only affects *SLCO1B1* (start ≥ 21.1 Mb; Figure S7) exhibited a significantly milder increase in total bilirubin (two-sided t-test: *p* = 3.1×10^−4^; Figure 5A), illustrating how mutations believed to be pathogenic in a digenic recessive framework can contribute to subtle changes in disease-associated phenotypes when present in an isolated heterozygous state.

**Figure 5.**
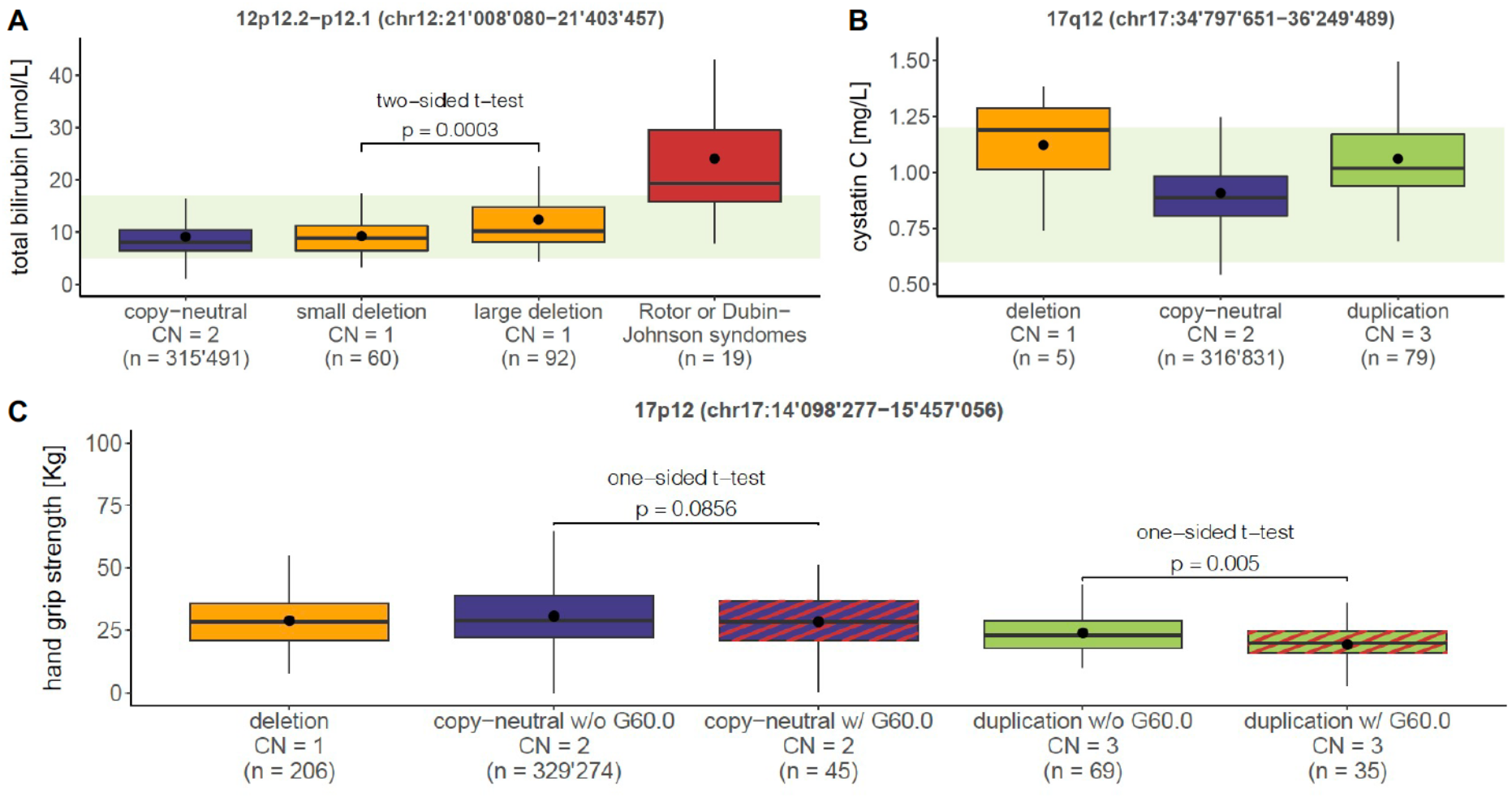
CNV-GWAS associations at Mendelian disorder loci. Boxplots showing levels of **(A)** total bilirubin in copy-neutral individuals, small or large 12p12.2-p12.1 deletion carriers, and Rotor/Dubin-Johnson syndrome patients (ICD-10 E80.6), **(B)** cystatin C in individuals with 17q12 deletion, copy-neutrality, or duplication, and **(C)** hand grip strength in individuals with 17p12 deletion, copy-neutrality, or duplication, split according to the presence (w/) or absence (w/o) of a neuropathy (ICD-10 G60.0; Red stripes). CNVR coordinates are at the top; Copy number (CN) and sample size are reported for each category; Dots show the mean; Outliers are not shown; Green bands show normal clinical range for **(A)** total bilirubin: 5-17 μmol/L and **(B)** cystatin C: 0.6-1.2 mg/L.

A second example is the association between the 1.5 Mb-long 17q12 duplication (chr17:34’797’651-36’249’489) and increased levels of kidney damage biomarkers, including cystatin C (β_dup_ = 0.15 mg/L, *p =* 4.2×10^−17^; Figure 5B), serum creatinine (SCr; β_dup_ = 13.0 µmol/L, *p =* 2.7×10^−16^; Figure S8A), and serum urea (β_dup_ = 0.93 mmol/L, *p =* 9.1×10^−10^; Figure S8B), as well as the inflammation biomarker C-reactive protein (CRP; β_mirror_ = 2.3 mg/L, *p =* 1.1×10^−6^; Figure S8C). Deletion of this interval is linked to renal cysts and diabetes syndrome (RCAD; MIM: 137920), a highly pathogenic and penetrant autosomal dominant disorder that can also be caused by mutations in the CNVR-overlapping gene *HNF1B* (Figure S8D). From a clinical perspective, RCAD is characterized by heterogenous structural and/or functional renal defects, neuro-developmental/psychiatric disorders, and maturity-onset diabetes of the young (MODY) (Mitchel et al., 2020). Due to the small number of deletion carriers (n = 6, regardless of phenotypic data availability) the deletion’s effect was not assessed by CNV-GWAS but the elevated levels of cystatin C (Figure 5B), SCr (Figure S8A), and urea (Figure S8B) in these individuals align with RCAD’s clinical description. Conversely, penetrance of the reciprocal duplication remains debated and only ∼20% of diagnosed patients report renal abnormalities (Mefford, 2021). Agreeing with a lower pathogenicity, we detected 16x more duplication than deletion carriers. Still, these individuals showed strong alterations in kidney function biomarkers (Figure 5B; Figure S8), suggesting tight gene dosage control on *HNF1B*.

Third, we zoomed in on the 1.4 Mb-long 17p12 duplication (Figure S9A) known as the main etiology of Charcot-Marie-Tooth (CMT) type 1A (MIM:118220), a demyelinating neuropathy of the peripheral nervous system characterized by progressive muscle wasting (Paassen et al., 2014). Correspondingly, duplication carriers showed decreased hand grip strength (chr17:14’098’277-15’457’056; β_dup_ = −9.8 Kg, *p =* 4.1×10^−39^; Figure 5C) and low SCr (chr17:14’098’277-15’468’444; β_dup_ = −13.8 µmol/L, *p =* 6.5×10^−18^; Figure S9B; EstBB: β_dup_ = −94.1 µmol/L, *p =* 5.2×10^−4^; Figure 3), indicating decreased muscle mass (Horowitz and Staros, 2019). We next sought to identify the proportion of duplication carriers (regardless of phenotypic data availability) diagnosed with CMT or related hereditary motor and sensory neuropathies and detected 48 and 38 diagnoses among the 331’206 copy-neutral individuals and 107 duplication carriers, respectively. While there is a clear enrichment for CMT diagnoses among duplication carriers (Fisher’s exact test: odds ratio = 3’668, *p* < 2.2×10^−16^), only 36% of duplication carriers were clinically identified. To test whether these individuals presented with more extreme clinical manifestations, we compared grip strength and SCr levels in duplication carriers with or without a neuropathy diagnosis and observed that while the former group exhibited lower grip strength (one-sided t-test: *p* = 0.005; Figure 5C), no difference was detected in SCr levels (one-sided t-test: *p* = 0.384; Figure S9B). Importantly, there was no age difference between diagnosed (mean = 55.5 years) and undiagnosed (mean = 56.2 years) duplication carriers (two-sided t-test: *p* = 0.650), indicating that results do not reflect biases regarding age of disease onset.

Through these examples, we show that well-established pathogenic CNVs can modulate disease-associated phenotypes in the general population without necessarily causing clinically diagnosable disorders, supporting a model of variable expressivity for the involved genetic loci (Chen et al., 2016; Cooper et al., 2013; Goodrich et al., 2021; Wright et al., 2019).

### CNV-GWAS signals suggest new gene functionalities

CNV-GWAS signals can be used to validate or generate hypotheses regarding the function of encompassed genes, as shown with the association between the CN of a 1.2 Mb 16p13.11 DNA stretch (chr16:15’120’501-16’308’285) and female reproductive traits. Specifically, duplication of the region correlated with decreased age at menarche (β_dup_ = −0.6 years, *p =* 2.0×10^−10^) and menopause (β_dup_ = −1.8 years, *p =* 1.7×10^−6^), whereas its deletion correlated with increased age at menarche (β_del_ = 1.1 years, *p =* 3.6×10^−7^), suggesting a shift in reproductive timing associated with the region’s CN (Figure 6A-B) that aligns with a low, albeit positive genetic correlation between the two traits (Neale Lab UKBB genetic correlation; see Web Resources). The effect of the duplication on age at menarche (β_dup_ = −0.6 years, *p =* 1.8×10^−2^) and menopause (β_dup_ = −2.6 years, *p =* 4.5×10^−2^) were confirmed with nominal significance in the EstBB (Figure 3A) and a SNP-GWAS signal for age at menarche (rs153793) colocalized with the CNVR (Day et al., 2017; Figure 6C). Literature supports the role of *MARF1* in the observed association. First, *MARF1* (o/e = 0.05 [0.03-0.12]; pLI = 1) and *MYH11* (o/e = 0.22 [0.16-0.30]; pLI = 0.77) are the only encompassed genes under evolutionary constraint according to GnomAD (upper bound of observed/expected ratio (o/e) < 0.35; Figure 6C; Table S7) (Karczewski et al., 2020; see Web Resources). Second, *MARF1* was shown to play an essential role in murine oogenesis by fostering successful completion of meiosis and cytoplasmic maturation, as well as through protection of germline genomic integrity (Su et al., 2012a). The gene’s function was further supported by studies in fly (Kawaguchi et al., 2020) and goat (Islam et al., 2019), as well as by two human case reports of females with *MARF1* mutations and reproductive phenotypes (Katari et al., 2018; Yang et al., 2018). Importantly, the female-specific role of *MARF1* (Islam et al., 2019; Katari et al., 2018; Kawaguchi et al., 2020; Su et al., 2012a, 2012b; Yang et al., 2018) aligns with the absence of association between the CNVR and our proxies for male sexual maturation (*i*.*e*. age at first facial hair and balding). Although further investigations are warranted to characterize the function of *MARF1* in human female reproduction and assess the role of other genes and regulatory elements within the CNVR, this illustrates how CNV-GWAS data can be leveraged to generate plausible hypotheses regarding new gene functionalities.

**Figure 6.**
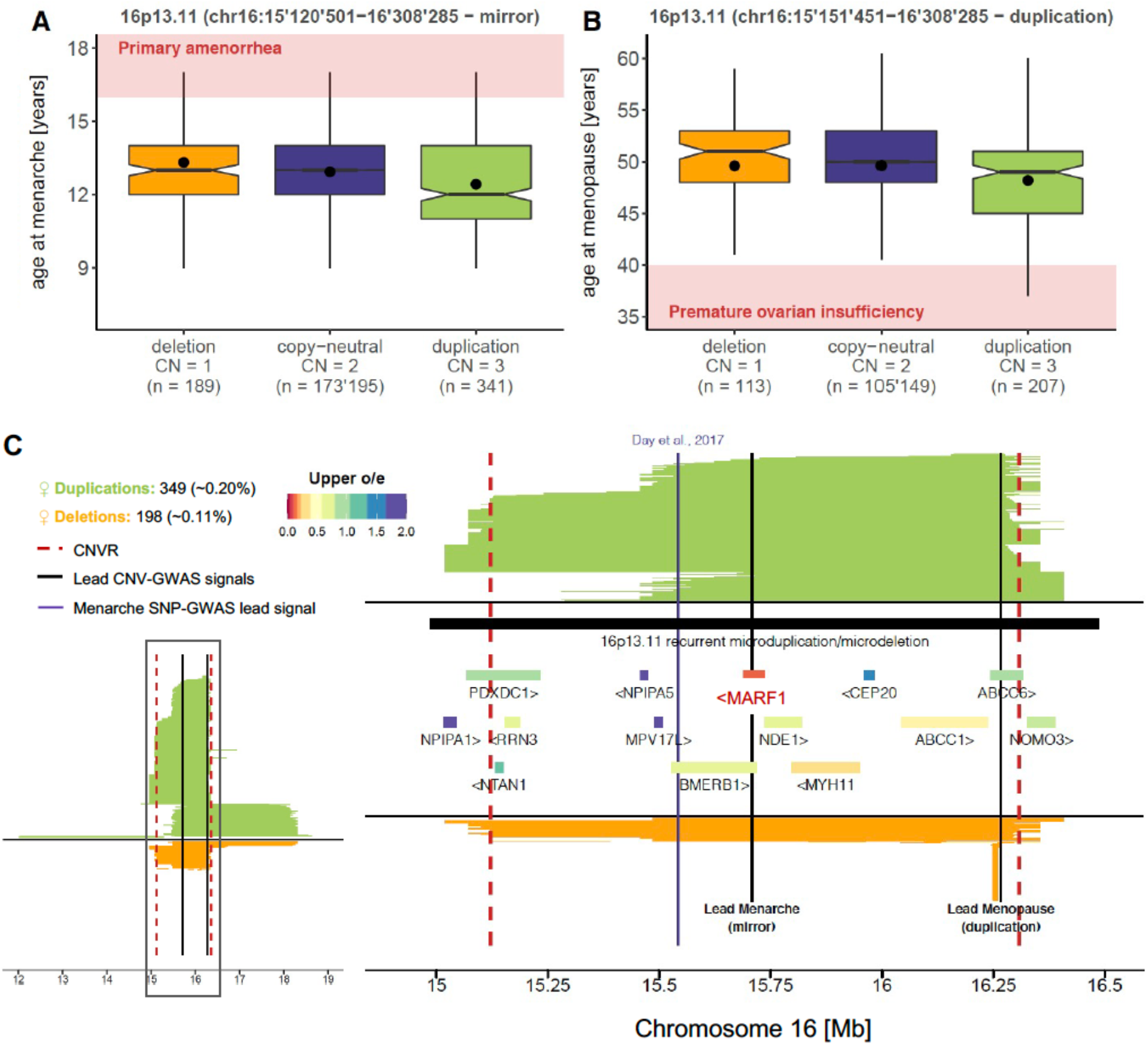
*MARF1* as a putative gene involved in human female reproduction. Boxplots representing age at **(A)** menarche and **(B)** menopause in individuals with 16p13.11 deletion, copy-neutrality, or duplication. CNVR coordinates are at the top; Copy number (CN) and sample size are reported for each category; Dots show the mean; Notches represent median ± 1.58*IQR/√n; Outliers are not shown; Red bands indicate pathogenic values corresponding to **(A)** primary amenorrhea (age at menarche > 16 years) (Gasner and Rehman, 2021) and **(B)** premature ovarian insufficiency (age at menopause < 40 years) (Walker and Tobler, 2021). **(C)** Mapping of CNVs overlapping the 16p13.11 CNVR. Number and frequency of duplications and deletions are at the top left; Left plot shows all overlapping CNVs and right plot focuses on the associated CNVR represented with red dashed lines (chr16:15’120’501-16’308’285); Duplications are in green, deletions in orange; Black lines indicate the location of the lead signal for age at menarche (mirror) and menopause (duplication-only); Purple line indicates the SNP associated with age at menarche; Overlapping recurrent CNVR is shown in black and protein-coding genes are colored according to the upper bound of the confidence interval for the observed/expected (o/e) mutation ratio in the GnomAD database.

### The deleterious impact of a high CNV burden

Moving beyond single CNVs, we estimated the impact of an individual’s total CNV burden on complex traits. Each participant’s autosomal CNV (duplication and deletion), duplication, and deletion burden was calculated as the number of Mb or genes rearranged by CNVs. Both Mb and gene burden metrices correlated well (*ρ*: 0.71-0.74) and while we observed high correlations (*ρ:* 0.40-0.92) between the CNV and duplication/deletion burdens, duplication and deletion burdens were uncorrelated (Figure 7A). Thirty-five of the fifty-seven traits analyzed by CNV-GWAS significantly associated with at least one burden metric (*p* ≤ 0.05/63 = 7.9×10^−4^, see Methods), often showcasing negative health consequences such as increased levels of adiposity, liver or kidney damage biomarkers, leukocytes, glycemic values, or anxiety and decreased global physical capacity or intelligence (Figure 7B; Table S8). Harmful phenotypic consequences were often best captured by the number of deleted genes, in line with a higher prevalence of sensitivity to decreased (*i*.*e*. haploinsufficiency), as opposed to increased (*i*.*e*. triplosensitivity) gene dosage (Collins et al., 2021).

**Figure 7.**
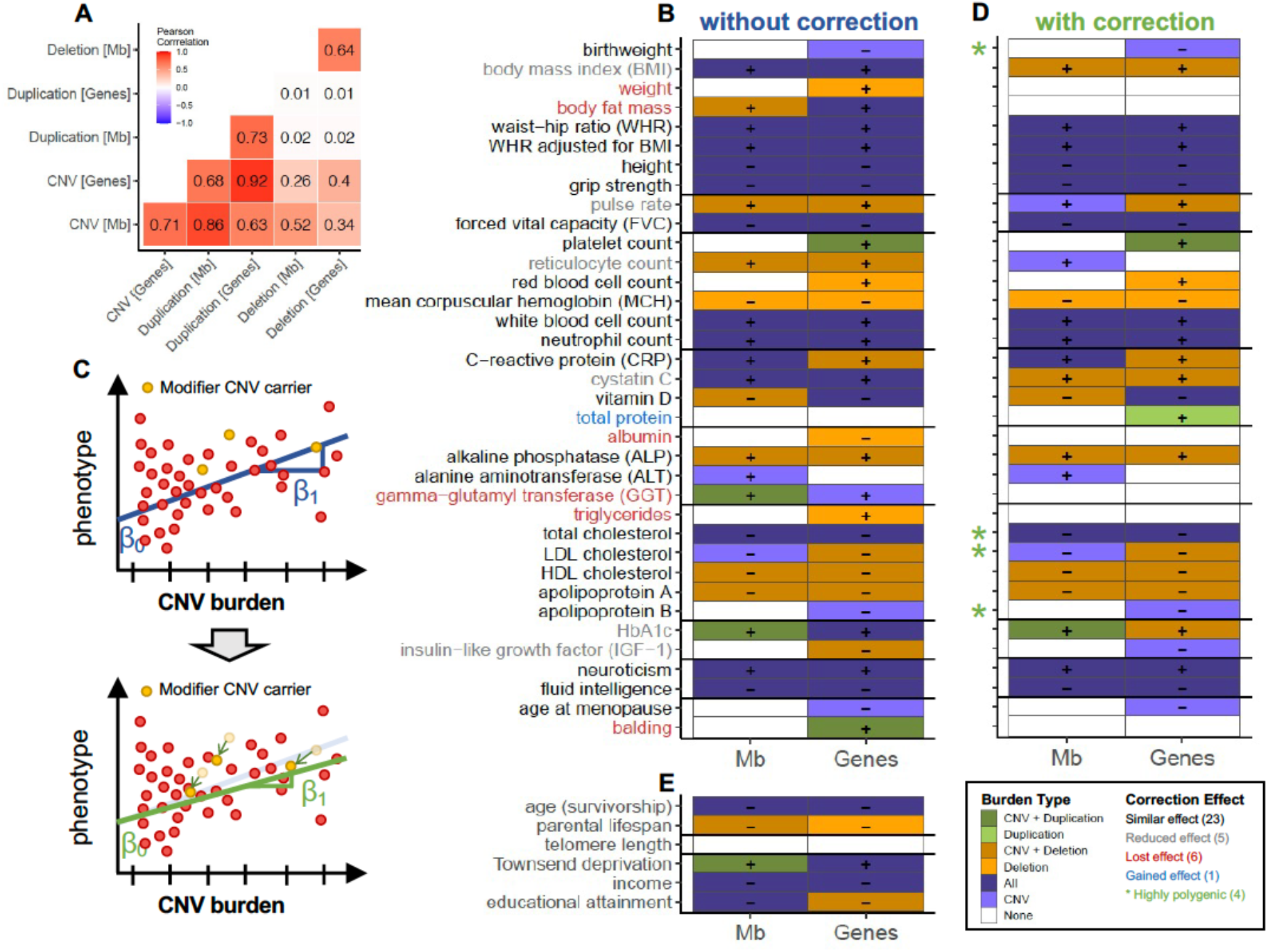
The negative impact of the CNV burden on complex traits. **(A)** Pearson correlation across the 6 burden metrices. **(B)** Significant associations (*p* ≤ 0.05/63 = 7.9×10^−4^) between the CNV burden, expressed as the number of Mb or genes affected by CNVs (x-axis) and phenotypes assessed through CNV-GWAS (y-axis). Color represents the type of burden (*i*.*e*. CNV, duplication, deletion, or a combination thereof) that was found to increase (“+”) or decrease (“-”) the considered phenotype. **(C)** Schematic representation of the correction for modifier CNVs: individuals carrying a CNV significantly impacting the trait under scrutiny are identified (top). Both phenotype and burden are corrected for the modifier (green arrows; bottom) and a new linear regression is fitted on the corrected data (bottom). **(D)** Significant associations (*p* ≤ 0.05/63 = 7.9×10^−4^) between the CNV burden after correction for modifier CNVs. Phenotype label color indicates whether the association between the CNV burden and the trait was lost (red), decreased (grey), identical (black), or strengthened (blue) after the correction. Green stars mark highly polygenic traits that associate with the CNV burden without having any significant CNVR association. **(E)** Significant associations (*p* ≤ 0.05/63 = 7.9×10^−4^) between the CNV burden and life history traits (y-axis). **(D-E)** follow the legend in **(B)**.

In a second step, we corrected each individual’s phenotype and burden for the presence of CNVs found to be GW-significantly associated with the same trait and performed the burden analyses anew to ensure that associations were not solely driven by CNV-GWAS uncovered loci (Figure 7C; Table S8). Whereas the association was lost for albumin, balding, body fat mass, GGT, triglycerides, and weight, indicating a mono-or oligogenic CNV-architecture, 30 traits remained associated with the burden. Specifically, 23 traits remained similarly associated, 6 associations persisted despite a decreased number of associated metrices, and one trait (total protein) gained in association strength (Figure 7D). Among these, birth weight, total cholesterol, low-density lipoprotein (LDL) cholesterol, and apolipoprotein B were significantly associated with the burden (Figure 7D), without any particular CNVR impacting them (Figure 2D). Overall, this reveals that as established for SNPs (Canela-Xandri et al., 2018; Visscher et al., 2017; Watanabe et al., 2019), the CNV architecture underlying most complex traits is highly polygenic, furthermore suggesting the presence of additional associations which we currently lack the power to detect.

The CNV burden extended its impact to global aspects of an individual’s life, illustrated through a negative correlation with several socio-economic factors, including decreased educational attainment (EA; β_burden_ = −0.07 years/Mb, *p* = 4.4×10^−11^) and income (β_burden_ = −1’593 £/year/Mb, *p* = 2.9×10^−60^), and increased Townsend deprivation index (β_burden_ = 0.04 SD/Mb, *p* = 3.6×10^−7^) (Figure 7E; Table S9). While we did not observe any effect on age- and sex-corrected telomere length, an individual’s CNV burden negatively associated with both parental lifespan (β_burden_ = −0.21 years/Mb, *p* = 1.4×10^−5^) and age (survivorship proxy; β_burden_ = −0.18 years/Mb, *p* = 1.1×10^−7^), suggesting that the deleterious impact of CNVs contributes to decreased longevity (Figure 7E; Table S9). Given this, we questioned whether the CNV burden was transmitted across generations at a Mendelian rate. Taking advantage of the presence of a sibling in the UKBB for 16’179 individuals assessed in our previous analyses, we calculated that the average fraction of CNVs these individuals share with their siblings was 27%. While substantially higher than for random pairs (0.7%), it only represents 54% of the expected fraction of shared additive genetic variance among siblings (50%) (Fisher, 1918). Together, these results describe the broadly deleterious impact of CNVs on a wide range of complex traits in the general population and suggest that most traits are influenced by a polygenic CNV-architecture.

## DISCUSSION

By coupling CNV calls to the rich phenotypic data available in the UKBB, we generated a roadmap of clinically relevant CNV-trait associations that allowed us to gain deeper and novel insights into specific biological pathways and put forward general patterns describing the role of CNVs on complex human traits in the general population.

Our UKBB CNV landscape matched previous reports (Aguirre et al., 2019) and while many of the 131 CNV-GWAS signals overlapped previously reported associations (Aguirre et al., 2019; Macé et al., 2017; Owen et al., 2018; Sinnott-Armstrong et al., 2021), our analyses shed light on several examples that had not been extensively studied in population-based cohorts, such as the association between the CN of the *MARF1*-encompassing 16p13.11 CNVR and female reproductive phenotypes. Another example is the association between the highly frequent *RHD* deletion and increased reticulocyte count and decreased platelet count and HbA1c levels. If observed effects are mild, lack or strongly reduced expression of all Rh antigens, a rare condition referred to as Rh deficiency or Rh_null_ syndrome (MIM: 617970, 268150), is associated with increased erythrocyte osmotic fragility, which results in hemolytic anemia (Nash and Shojania, 1987). Hemolytic anemia is characterized by increased reticulocyte count (Rai et al., 2021) and was shown to falsely lower HbA1c levels due to the decreased erythrocyte lifespan (Goldstein et al., 1995), putting forward the hypothesis that heterozygous deletion of *RHD* leads to subclinical tendencies towards hemolytic anemia. Similar trends were observed in individuals with a Rh^-^ blood type, which can be caused by various polymorphisms (Avent and Reid, 2000). Of note, the latter cohort consisted of pregnant women and reticulocyte and platelet counts have been reported to increase and decrease, respectively, along pregnancy (Akinlaja, 2016). Hence, despite correcting for pregnancy status and gestational weeks, interactions between the Rh^-^ blood group and pregnancy cannot be excluded. While the impact of Rh blood type on hematological traits awaits validation, these examples illustrate how CNV-GWAS signals can be leveraged to gain new biological insights.

Through the combined use of three association models, our CNV-GWASs revealed general patterns through which CNVs modulate phenotypes. Geared towards the discovery of mirror effects, several such cases were observed. More difficult to detect with our analytical framework, U-shape effects were also witnessed, as illustrated with the 17q12 reciprocal CNV, which increases kidney damage biomarkers. Together, this confirms the existence of different mechanisms through which altered dosage can influence phenotypes. We further provide evidence for a broad and nuanced role of CNVs in shaping complex traits, as both common (frequency ≥ 1%) and rare (frequency < 1%) CNVs mapping to regions previously involved by SNP-GWAS contribute to phenotypic variability in the general population, with rare CNVs having larger effects sizes than common ones. In contrast, many signals mapped to regions involved in Mendelian disorders. Matching the increasing awareness around variable penetrance and expressivity of these loci (Chen et al., 2016; Cooper et al., 2013; Fahed et al., 2020; Goodrich et al., 2021; Oetjens et al., 2019; Wright et al., 2019), we show that CNVs overlapping these regions can impact disease-associated phenotypes in the general population without necessarily causing clinically diagnosable disorders. In line with the allelic series observed for *SLCO1B1* (Sinnott-Armstrong et al., 2021), our data show that the heterozygous deletion of one, as compared to both of the genes involved in Rotor syndrome leads to a gradual, non-pathogenic increase in total bilirubin, which remains below the values observed in homozygously affected individuals. In another example, we show that while individuals carrying the 17p12 CMT1A duplication show signs of muscle wasting, in agreement with the disease’s clinical description (Paassen et al., 2014), only 36% of the most severely affected duplication carriers were diagnosed, consistent with a variable expressivity model of the genotype. By studying CNVs in the general population, as opposed to clinical cohorts selected based on phenotypic criteria or family history, it becomes possible to re-assess the frequency, penetrance, expressivity, and inheritance pattern or pathogenic CNVs, providing a more complex and nuanced – but also broader – understanding of their phenotypic impact.

Confirming the deleterious influence of a high CNV load on anthropometric traits (Dauber et al., 2011; Macé et al., 2017; Wheeler et al., 2013) and EA (Kendall et al., 2017; Männik et al., 2015; Saarentaus et al., 2021), we extended this observation to 36 additional traits, including socio-economic factors and global health biomarkers. Hypothesizing a link between telomere length and CNV formation, we tested whether the trait associated with the CNV burden but did not find any association. In contrast, telomere length specifically associated with the *BRCA1* cancer locus (Miki et al., 1994) (chr17:41’197’733-41’258’551, β_dup_ = 0.45 SD, *p =* 1.9×10^−8^; N_dup_ = 154; N_del_ = 17), paralleling findings that long telomere-associated SNPs also associate with increased cancer risk (McNally et al., 2019), possibly by mediating replicative senescence escape. Finally, given the low number of deceased UKBB participants, we used proxies to assess whether a high CNV burden affected an individual’s lifespan. We found that individuals with advanced age had a lower CNV burden, matching the depletion of life-shortening alleles in older UKBB participants (McDaid et al., 2017) and suggesting improved health/decreased mortality in individuals with a low CNV load. Parental lifespan negatively correlated with the CNV burden. While lower than the expected, a substantial fraction of CNVs (27%) was shared among siblings and thus inherited from either parent. Several factors can contribute to decreased CNV-sharing: First, high false negative rates in microarray-based CNV calls might impair detection of shared CNVs in the sibling. Second, selection bias during gametogenesis and/or early development or ascertainment bias (*i*.*e*. increased participation rate in healthier siblings) might eliminate siblings carrying deleterious CNVs. Finally, *de novo* CNVs could decrease CNV-sharing between siblings. Unfortunately, confounders - such as CNV length, which affects both detection capacity and pathogenicity – prevent the assessment of these factors separately. Nevertheless, the lower-than-expected CNV inheritance allow speculating that an even stronger signal would be obtained providing access to high-quality parental CNV genotypes. While further studies are required to confirm the life-shortening effect of a high CNV load, our data clearly illustrates the deleterious impact of CNVs on an individual’s global health.

Lastly, combining results from the CNV-GWAS and burden analyses allowed us to gain an understanding of the CNV architecture underlying studied traits. Many loci highlighted by the CNV-GWAS represent well-known recurrent CNVRs. However, due to the difficulty to gather large cohorts of individuals with rare genomic events, complete phenotypic characterization of most of these loci is still missing and limited to easily assessed anthropometric traits or severely debilitating neuro-developmental/-psychiatric disorders. Our results provide a map of the pleiotropic consequences of these CNVR on over 50 medically relevant traits. Some of these associations might be mediated through primary phenotypic changes. For instance, it is known that obesity causally alters blood pressure, serum lipids, glycemic values, and liver biomarkers (Fall et al., 2013; Freathy et al., 2008; Würtz et al., 2014). Importantly, most CNVRs are large and gene-rich, potentially harboring several causal genes. One of the next challenges will be to untangle primary from secondary associated traits and narrow down causal region(s) in pleiotropic multi-genic CNVRs. Importantly, the substantial overlap between CNV- and SNP-GWAS signals speaks for the presence of shared genetic mechanisms and indicates that both mutational classes can be exploited synergistically to pinpoint causal genes and elucidate their biological function. In parallel, we observed a high degree of CNV-polygenicity, as 29 out of 35 traits remained associated with the CNV burden after correction for modifier CNVRs. For albumin, balding, body fat mass, GGT, triglycerides, and weight, signals from the CNV-GWAS capture the bulk of phenotypic variability caused by CNVs. At the other end of the spectrum, apolipoprotein B, birth weight, LDL cholesterol, and total cholesterol were solely associated with the CNV burden, indicating either a polygenic CNV-architecture and/or the presence of rare (frequency ≤ 0.005%) high impact CNVs that were not assessed by CNV-GWAS. Among these traits, decreased birth weight, which associated with a high CNV burden, has been linked to increased risk for metabolic syndrome, obesity, and various other diseases in adulthood (Armengaud et al., 2021; Barker et al., 1993), opening the question as to whether some of the deleterious effects of the CNV burden are rooted in early development. Strikingly, the three other traits relate to plasma lipids, contrasting with the few CNV-GWAS signals detected for these traits (16p11.2 BP2-BP3 – HDL cholesterol and apolipoprotein A; 16p11.2 BP4-BP5–triglycerides). Speaking for the high polygenicity of plasma lipids, GWAS on 35 blood biomarkers in the UKBB found an average of 87 versus 478 associations/trait for non-lipid, as compared to lipid traits (Sinnott-Armstrong et al., 2021), motivating further research to elucidate the CNV architecture of these phenotypes. Collectively, these results illustrate a more complex than expected contribution of CNVs in shaping the genetic architecture of complex human traits.

When interpreting results, it is important to keep in mind the limitations of the current study. First, CNVs were called based on microarray data. In addition to high false positive rates associated to array-based CNV calls, this renders the study blind to variants in regions not covered by the array and limits resolution – both in absolute length and exact break point location – of the assessed CNVs (Valsesia et al., 2013). To mitigate these issues, we used a pipeline to filter CNVs and transform calls to the probe level (Macé et al., 2016, 2017). Future release of large whole-genome sequencing datasets combined to progress in CNV detection tools should resolve these issues and lead to the discovery of new small-scale CNV-trait associations (Chen et al., 2021; Halvorsen et al., 2020). Second, as the aim of this study was to provide a set of trustworthy CNV-trait pairs for follow-up analyses, we used stringent criteria both for CNV filtering and association detection, at risk of missing some true associations. Following previous works (Aguirre et al., 2019; Macé et al., 2017), we omitted to account for the number of assessed phenotypes through CNV-GWAS. Even with an extremely stringent experiment-wide threshold for significance (*p* ≤ 0.05/(11’804*57) = 7.4×10^−8^), 68 out of 131 (52%) CNV-GWAS signals, spanning 33 traits, remained significantly associated. As most signals selected for follow-up analyses met this threshold, at least 24% of the signals that did not are supported by a SNP-GWAS signal, speaking for the fact that these are true positives. Third, while we tried to reproduce UKBB discoveries in the EstBB, there are only few cohorts with sufficiently large genetic and phenotypic coverage to replicate original finding at adequate power, meaning we had to rely on literature evidence to gauge the validity of our results. Fourth, despite showing substantial colocalization of CNV- and SNP-GWAS signals, we did not assess whether trait-associated CNVR were robustly enriched for SNP-GWAS signals. Due to the non-random genomic distribution and complex nature of CNVs, simulating the null scenario goes beyond the scope of this paper. Of note, the number of SNP-/CNV-signal colocalizations is likely to be underestimated as manual literature search revealed additional examples that were missed by our annotation pipeline, such as the SNP-GWAS signal overlapping the 16p13.11 CNVR (Day et al., 2017). Fifth, the present study is limited to individuals of white British ancestry. As CNV frequencies vary across populations (Campbell et al., 2011; Chen et al., 2011; Li et al., 2009; Sudmant et al., 2015), extending this approach to additional ancestral groups is likely to result in the discovery of additional associations. Finally, the UKBB suffers from a “healthy cohort” bias (Fry et al., 2017). As our primary focus was to assess the impact of CNVs in a healthy population, this bias can be used at our advantage, as it incorporates CNV carriers with sub-clinical phenotypes to the studied cohort, providing a lower bound for effect size estimates (Goodrich et al., 2021; Martin et al., 2020; Wright et al., 2019). However, this means that the cohort is depleted for severely affected cases. As ≤ 0.005% participants carried the highly pathogenic 17q12 (RCAD) or 22q11.21 (DiGeorge syndrome) deletions, these associations were not tested, even though we show that 17q12 deletion carriers present abnormal levels of renal biomarkers. Extending the analysis to low frequency/high impact CNVRs would allow to better distinguish between CNVR mechanisms of action – with the remaining caveat the effects will be underestimated due to selection bias - and will be the focus of future work.

In conclusion, our study provides a map of high-confidence CNV-trait associations. While we started to explore some of the reported signals, collective efforts will be required to validate and interpret the bulk of these discoveries and we hope that this resource will be useful for researchers and clinicians aiming at improving the characterization of recurrent CNVs. Furthermore, our study revealed the nuanced role of CNVs along the rare versus common disease spectrum, their shared mechanisms with SNPs, as well as a wide-spread polygenic CNV-architecture, consolidating the growing body of evidence implicating CNVs in the shaping of complex human traits.

## MATERIAL AND METHODS

### 1. Study material

#### The UK Biobank cohort

The UK Biobank (UKBB) is a volunteer-based cohort of ∼500’000 individuals (54% females) from the general UK population. Individuals were aged between 40-69 years at recruitment and underwent microarray-based genotyping, as well as extensive phenotyping, which is constantly extended and includes physical measurements, blood biomarker analyses, socio-demographic and health-related questionaries, as well as linkage to medical health records (Bycroft et al., 2018). Participants signed a broad informed consent form and data was accessed through the application number 16389.

#### The Estonian Biobank cohort

The Estonian Biobank (EstBB) is a population-based cohort encompassing ∼20% of Estonia’s adult population (∼200’000 individuals; 66% females; https://genomics.ut.ee/en/research/estonian-biobank). Individuals underwent microarray-based genotyping at the Core Genotyping Lab of the Institute of Genomics, University of Tartu and general data, including basic body measurements, were collected at recruitment. Project-based questionnaires were sent later and filled on a voluntary basis. Health records are regularly updated through linkage with the national Health Insurance Fond and other relevant databases, providing sporadic access to blood biomarker measurements and medical diagnoses (Leitsalu et al., 2015). All participants signed a broad informed consent form and analyses were carried out under ethical approval 1.1-12/624 from the Estonian Committee on Bioethics and Human Research and data release N05 from the EstBB.

#### The CHUV maternity cohort

The Lausanne University Hospital (CHUV) maternity cohort aims to study maternal health. Initially designed as a serological surveillance study of maternal toxoplasmosis infections, approval from the Ethics Committee of Vaud (CER-VD) was obtained for data reusage under the project ID 2019-00280 to investigate maternal and fetal outcomes. Rh blood group was serologically determined for 5’164 women. Reticulocyte count, platelet count, HbA1c levels, intrapartum reports, and International Classification of Diseases, 10^th^ Revision (ICD-10) codes were sporadically collected between 2009-2014.

#### Software versions

Copy number variants (CNVs) were called with PennCNV v1.0.5 (Wang et al., 2007), using the PennCNV-Affy package (27/08/2009); CNV were filtered based on a quality scoring pipeline (Macé et al., 2016); Various genetic analyses were conducted with PLINK v1.9 and PLINK v2 (Chang et al., 2015), as indicated in text; Genome annotation was performed with ANNOVAR (24/10/2019) (Wang et al., 2010); Meta-analysis was carried out with GWAMA v2.2.2 (Mägi and Morris, 2010); Statistical analyses were performed with R v3.6.1 and graphs were generated with R v4.0.3.

### 2. The CNV landscape of the UKBB

#### Genotype data

Details of genotype data acquisition and quality control (QC) have been previously described (Bycroft et al., 2018). Briefly, UKBB participants were genotyped on two similar arrays (95% probe overlap): 438’427 samples (95 batches) were genotyped with the Applied Biosystems UK Biobank Axiom Array (825’927 probes) and 49’950 samples (11 batches) were genotyped with the Applied Biosystems UK BiLEVE Axiom Array by Affymetrix (807’411 probes). All results in this study are based on the human reference build GRCh37/hg19.

#### Sample selection

Related, gender mismatched, high missingness, non-white British ancestry, and retracted samples were excluded (*used*.*in*.*pca*.*calculation* = 0 and *in*.*white*.*British*.*ancestry*.*subset* = 0 in Sample-QC v2 file). To protect the analysis from somatic chromosomal aberrations, individuals with self-reported (#20001, codes: 1047, 1048, 1050, 1051, 1052, 1053, 1055, 1056, 1056; UKBB update 03/2020) and/or hospital diagnosed (#41270; all ICD-10 codes mapping to *cancer of lymphatic and hematopoietic tissue*’s exclusion range in the PheCode Map 1.2 (beta), accessed 09/12/2020 (Wu et al., 2019); UKBB update 08/2019) blood malignancy were excluded. CNV outliers were later removed (see *CNV quality score*). All reported results are for 331’522 unrelated white British UKBB participants (54% females).

#### CNV calling

CNVs on chr1-22 and pseudoautosomal regions, hereafter referred to as *autosomal CNVs*, were called in parallel for the 106 UKBB genotyping batches using PennCNV. Individual intensity files were generated from the B allele frequency (BAF) and Log R Ratio (LRR) files available on the UKBB portal and missing values (−1) were set to NA. Batch-specific population frequency of the B allele (PFB) files were generated by computing the batch’s mean BAF. Probes with missing PFB were removed in a batch-specific way. The hidden Markov Model file for Affymetrix genome-wide 6.0 array was downloaded as part of the PennCNV-Affy package and used without training. The GC model file was generated following instructions of `cal_gc_snp.pl`, using the gc5Base file (03/2020) downloaded from the UCSC Genome Browser (https://hgdownload.cse.ucsc.edu/goldenPath/hg19/database/). Above-described files were used to call CNVs with confidence score using `detect_cnv.pl` with genomic wave adjustment. Adjacent CNVs (gap ≤ 20% of merged CNV length) were merged with `clean_cnv.pl`. ChrX CNVs were called separately with the arguments `-chrx` and `-sexfile` when running `detect_cnv.pl,` copy-neutral losses of heterozygosity resulting from male chrX hemizygosity were excluded, and adjacent CNVs were merged.

#### CNV Quality Score

Hurdles linked to CNV analysis include high false positive rates and variability in break points. To mitigate these issues, a post-PennCNV processing pipeline was used to attribute a quality score (QS) to each CNV and transform calls to the probe level (Macé et al., 2016, 2017). QSs range from −1 to 1 and reflect the probability of a CNV to be a true positive (−1 = likely deletion; 1 = likely duplication; ∼0 = low confidence CNVs). PennCNV `filter_cnv.pl` was run separately on autosomal and chrX (with `-chrx`) CNVs, resulting in two sample-level QC files. These were combined by adding the number of chrX CNVs to the autosomal sample-level QC file. A single file containing all called CNVs, as well as associated CNV- and sample-level QC metadata, was generated. CNVs originating from samples genotyped on plates with a mean CNV count per sample > 100, as well as those originating from samples with > 200 CNVs or a single CNV > 10 Mb were excluded, as these might be indicative of batch-effects, genotyping errors, or extreme chromosomal abnormalities. A QS was calculated for each CNV and linear coordinates were transformed into a per-chromosome *probe × sample* matrix, with entries reflecting the QS attributed to the CNV mapping to these probes, 0 indicating copy-neutral probes. Individuals with no CNVs were added as all-0 columns.

#### Converting CNV calls into PLINK format

QS matrices were converted to PLINK binary file sets. Probes with ≥1 high confidence CNVs, stringently defined by |QS| ≥ 0.5, were retained and encoded into 3 file sets to accommodate analyses according to a mirror (PLINK_CNV_), duplication-only (PLINK_DUP_), or deletion-only (PLINK_DEL_) association model, using `--make-bed` (PLINK v1.9) (Table 1).

**Table 1.** Encoding of high-confidence CNVs (|QS| ≥ 0.5) from QS matrices into 3 PLINK file sets.

| Association Model | Mirror | Duplication-only | Deletion-only |
| --- | --- | --- | --- |
| PLINK file set | PLINK<sub>CNV</sub> | PLINK<sub>DUP</sub> | PLINK<sub>DEL</sub> |
| **Deletion** (QS < -0.5) | AA | 00 | TT |
| **Neutral** (-0.5 ≤ QS ≤ 0.5) | AT | AT | AT |
| **Duplication** (QS > 0.5) | TT | TT | 00 |

#### CNV frequency calculation

The genotype count `--freqx` function (PLINK v1.9) was applied to the 740’434 probes stored in PLINK_CNV_. Results were used to exclude 41’670 array-specific probes with genotype count missingness > 5% and calculate each probe’s 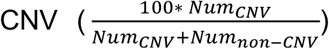, 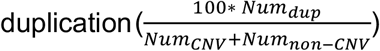, and 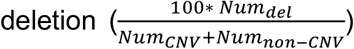 frequencies [%], with *Num*_*non-CNV*_, *Num*_*dup*_, and *Num*_*del*_, the number of individuals carrying 2, < 2, and > 2 copies of that probe, respectively, and *Num*_*CNV*_ = *Num*_*dup*_ + *Num*_*del*_.

### 3. CNV association studies in the UKBB

#### CNV probe selection and number of effective tests

Association studies were restricted to probes with a CNV, duplication, or deletion frequency ≥ 0.005% for the mirror, duplication-only, or deletion-only model, respectively. To group probes at the core region of CNVs while retaining variability at breakpoints, probes were pruned at r^2^ > 0.9999 by applying `–-indep-pairwise 500 250 0.9999` (PLINK v2) to PLINK_CNV_, PLINK_DUP_, and PLINK_DEL_. As these probes remained correlated, we determined the number of effective tests, *N*_*eff*_ (Gao et al., 2008). A per-chromosome *probe × sample* matrix *G* was generated, with genotypes taking values of −1 (deletion), 0 (copy-neutral), and 1 (duplication). Chromosome-wise *N*_*eff*_ were defined as the number of eigenvalues required to explain 99.5% of the variance in *G* and were summed up. *N*_*eff*_ was estimated at 11’804, setting the genome-wide (GW) threshold for significance at *p* ≤ 0.05/11’804 = 4.2×10^−6^. Accounting solely for duplications or deletions resulted in lower *N*_*eff*_ estimates but we used the same conservative threshold for all three association models.

#### Phenotype selection

Fifty-seven continuous traits were selected based on data availability and presumed high heritability. Fifty-four were defined as the mean of measured instances. Three were composite traits: Grip strength, as the mean of *hand grip strength left* and *right* (#46, #47); Waist-to-hip ratio (WHR), as the ratio between *waist* and *hip circumference* (#48, #49); WHR adjusted for body mass index (WHRadjBMI), by correcting WHR for BMI in a sex-specific fashion. Two were male-specific (*relative age of first facial hair* (#2375); *hair/balding pattern* (#2395)) and three were female-specific (*age when periods started (menarche)* (#2714); *age at menopause (last menstrual period)* (#3581); *birth weight of first child* (#2744)). Entries “do not know”, “only had twins”, “prefer not to answer” were set as missing. Traits were inverse normal transformed prior correction for sex (except for sex-specific traits), age, age^2^, genotyping batch, and PC1-40. Normal ranges were retrieved and converted from Symed MediCalc (see Web Resources).

#### Genome-wide copy number association studies (CNV-GWAS)

Associations between the copy number (CN) of selected probes and normalized covariate-corrected traits were performed at once using `–-glm` (PLINK v2) with the options `omit-ref`, `no-x-sex`, `hide-covar`, and `allow-covars`. For sex-specific traits, samples from the opposite sex were excluded. Three association models were applied: A mirror model, which assesses the additive effect of each additional copy of a probe, on PLINK_CNV_; A duplication-only model, which assess the impact of a duplication while disregarding deletions, on PLINK_DUP_; A deletion-only model, which assess the impact of a deletion while disregarding duplications, on PLINK_DEL_. Given the CNV encoding, effects were homogenized to “T” by multiplying β by −1 when A1 was “A”. GW-significant association (*p* ≤ 4.2×10^−6^) were retained.

#### Stepwise conditional analysis

Stepwise conditional analysis was performed on CNV-GWAS results to determine the number of independent signals per trait. For traits with ≥ 1 GW-significant signal, CNV information was extracted for the lead probe, with genotypes taking values of −1 (deletion), 0 (neutral), and 1 (duplication) when considering the mirror model and setting deletions or duplications to NA when considering the duplication-only or deletion-only models, respectively. Lead CNV probe effect was regressed out of the phenotype and association studies were conducted anew. These steps were repeated until no GW-associated probes were identified.

#### CNV region definition, merging, and annotation

CNV region (CNVR) boundaries were defined by the most distant probe within ≤ 3Mb and r^2^ ≥ 0.5 of each independent lead probe, using`–-show-tags` (PLINK v1.9) with options `--tag-kb 3000 --tag-r2 0.5` on the appropriated PLINK file sets. Signals from the different models were merged when the signal involved (1) the same trait, (2) overlapping CNVRs, and (3) were directionally concordant according to a mirror model. New CNVR boundaries were defined as the maximal CNVR and characteristics of the most significant model were retained for follow-up analyses. CNVRs were annotated with `annotate_variation.pl`, using hg19 RefSeq gene names `(--geneanno`; 08/06/2020) and NHGRI-EBI GWAS catalog associations (Buniello et al., 2019; `--regionanno`; 29/04/2021) from ANNOVAR.

### 4. Replication in the EstBB

#### Genotype data

Briefly, 202’282 EstBB participants were genotyped with Illumina GSAv1.0, GSAv2.0, GSAv2.0_ESTChip, and GSAv3.0_ESTChip2 arrays in 12 batches. Samples with genotype call rate < 98%, Hardy-Weinberg equilibrium test *p*-value < 1×10^−4^, or mismatched sex based on chrX heterozygosity were excluded and one of each duplicated sample was retained. Genotypes were re-clustered by manual realignment of cluster locations and intensity files (LRR and BAF) were created with Illumina GenomeStudio v2.0.4. A PFB file was generated from 1’000 randomly selected samples from batch 1. Only autosomal probes overlapping all GSA versions (excluding custom ESTChip probes) were carried over to the CNV detection step (671’035 probes; 242’091 probes overlap with UKBB).

#### Phenotype data

The 57 analyzed traits in the UKBB were queried in the EstBB database: 3 were available from measurements collected at enrollment (height, weight, BMI); 2 were collected using project-based questionnaires (age at menarche and menopause); 41 were retrieved from parsed notes in health registries; 11 did not have any corresponding term. Because most phenotypic measurements originate from health registries, they were gathered at different timepoints, by different practitioners, and were available for a limited subset of participants. In samples with repeated measurement for a given trait, the most recent one was retained. Traits with sample size ≥ 2’000 were inverse normal transformed prior correction for sex (except for sex-specific traits), age, age^2^, genotyping batch, and PC1-20.

#### CNV calling and copy number association studies

Autosomal CNVs were called by batch for 193,844 individuals, attributed a QS, and encoded into 3 PLINK binary file sets, following the procedure described for the UKBB. Samples originating from 2 batches with outlier genotyping intensity parameters, as well as genotyping plates with > 3 samples with either > 200 called CNV or a total length of CNV calls > 10 Mb were excluded. Individual samples meeting these criteria were further removed. Among related pairs (KING kinship coefficient > 0.0884), the sample with most available phenotypes was retained. CNV, duplication, and deletion frequencies among the 89’516 remaining samples were calculated for 671’035 probes and association studies were run as previously described for the UKBB, using probes with a CNV, duplication, or deletion frequency ≥ 0.005%, when running the mirror, duplication-only, or deletion-only association models, respectively. Using the most significant association model for the 131 merged UKBB signals, we selected the most significantly associated EstBB probe for the trait under investigation within the boundaries of the UKBB-defined CNVR. EstBB *p*-value were adjusted to account for directional concordance with UKBB effect sizes: In case of direction agreement, 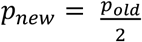, while in case of disagreement 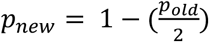,. Sufficient genomic variability and phenotypic data was available to assess replication of 61 out of 131 signals, setting the replication threshold for significance *p* ≤ 0.05/61 = 8.2×10^−4^.

#### Power analysis

Simulations were conducted to estimate the power of our replication study. We defined *β*_*i,j*_ as the standardized effect of probe *i* on trait *j* observed in the UKBB, *q*_*i,dup*_ and *q*_*i,del*_ the duplication and deletion frequencies of probe *i* in the EstBB, respectively, and *N*_*j*_ the sample size for trait *j* in the EstBB. When considering mirror signals, CNVs were simulated for *N*_*j*_ samples as:

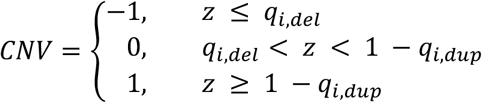

Where *z* follows *z* ∼ *U*(0,1). When considering duplication-only or deletion-only signals, duplications or deletions were simulated for *N*_*j*_ samples as *CNV* ∼ *Bernoulli*(*q*_*i,dup*_) or *CNV* ∼ *Bernoulli*(*q*_*i,del*_), respectively. Error terms *ε* for *N*_*j*_ samples were simulated according to a normal distribution *ε* ∼ *N*(0, *σ*^2^). For mirror signals, the noise variance *σ*^2^, was defined as 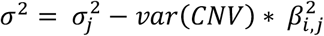, with 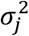 the observed standardized variance for trait *j* in the EstBB equaling 1. For duplication-only and deletion-only signals, *σ*^2^ was defined as 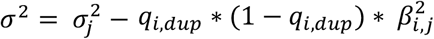 and 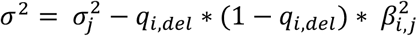, respectively. Phenotypes *Y* were simulated for *N*_*j*_ samples as *Y* = *CNV* * *β*_*i,j*_ + *ε*. When simulated data contained ≥ 1 CNV carrier, the *p*-value for the estimated effect size from the linear regression *Y* ∼ *CNV* was computed and retained. Otherwise, the *p*-value was set as missing. For each signal, 10’000 simulations were conducted and power was defined as the fraction of non-missing *p*-values ≤ 8.2×10^−4^. The expected number of replications, given available power, was estimated as the average power across assessed signals multiplied by the number of assessed signals.

### 5. Extended phenotypic assessment

#### Diseases diagnoses (ICD-10)

To assess patients’ disease status, ICD-10 diagnoses were used (#41270). For GGT-altering diseases, *heart failure* (I50), *malignant neoplasm of liver and intrahepatic bile ducts* (C22), gallbladder (C23), and *other unspecified parts of biliary tract* (C24), as well as other *diseases of the liver* (K70-K77) and the *gallbladder, biliary tract, or pancreas* (K80-K87) were considered. Rotor syndrome is classified with Dubin-Johnson syndrome under *other disorders of bilirubin metabolism* (E80.6). CMT is classified as *hereditary motor or sensory neuropathy* (G60.0), a diagnosis encompassing all forms of CMT, as well as other related neuropathies.

#### Lifestyle

To verify that the 22q11.23-GGT association was not confounded, we established a list of samples with high alcohol consumption as those with self-reported *daily or almost daily* alcohol intake (#1558), as well as those using GGT-increasing drugs (#20003, codes: 2038459704 (carbamazepine), 1140865426 (cimetidine), 1140909708 (furosemide), 1140869848 (methotrexate), 1140910706 (phenobarbital), 2038460076 (phenytoin)) (Dufour et al., 2000).

#### Life history

Burden analysis was extended to socio-economic factors including *Townsend deprivation index at recruitment* (#189), *average total household income before tax* (#738) averaged over measured instances (categories: £18’000, £24’500, £41’500, £76’000, and £100’000), and *age completed full time education* (#845), as a proxy for educational attainment (EA). Assessed life history traits include *age at recruitment*, as a proxy for survivorship (# 21022), *mother’s* (#3526) and *father’s* (#1807) *age at death* (meta-analyzed as parental lifespan), as well as *relative leucocyte telomere length adjusted for the influence of technical parameters* (#22191). Entries matching “do not know or “prefer not to answer” were set as missing. Traits were inverse normal transformed prior correction for sex, age, age^2^, genotyping batch, and PC1-40, except for *age at recruitment*, which was not corrected for age and age^2^

### 6. *RHD* and hematological traits

#### Transcriptome-wide Mendelian Randomization

Using univariable transcriptome-wide Mendelian randomization (TWMR) (Porcu et al., 2019), the causal effect of differential *RHD* and *RSRP1* expression on hematological traits was estimated based on independent (r^2^ < 0.01) genetic variants. Expression quantitative trait loci (eQTLs) were obtained from the eQTLGen consortium and included *cis*-eQTLs (FDR < 0.05, 2-cohort filter) for ∼16,900 transcripts (Võsa et al., 2018). GWAS effect sizes originate from Neale Lab UKBB summary statistics (see Web Resources). Prior to the analysis, exposure and outcome datasets were harmonized, standardized effect size estimates were obtained by dividing the z-scores by the square root of the sample size, and palindromic variants and variants with allele frequency difference > 5% between the two datasets were removed. To ascertain robustness, analyses were performed excluding rs55794721, which had an extreme effect on both exposures and outcomes.

#### Association between Rh blood group and hematological traits

Impact of Rh^-^ blood group on platelet count, reticulocyte count, and HbA1c was assessed in the CHUV maternity cohort through multivariate linear regression that incorporates the covariates: age at measurement, gestational week at measurement, whether the woman was pregnant at measurement (57.5% for reticulocyte count, 35.6% for platelet count, 23.4% for HbA1c), and whether the women had a child prior to the measurement (78.9% for reticulocyte count, 72.7% for platelet count, 96.7% for HbA1c). For women with multiple measurements, one was randomly selected, giving preference to measurements taken outside of pregnancy and excluding measurements taken during a pregnancy that resulted in stillbirth or multiple births. For measurements taken during pregnancy, gestational week at measurement was calculated from date and gestational age at delivery. When gestational age at delivery was missing (52.9%), mean gestational age at delivery of the cohort (39.13 weeks) was used. For measurements outside of pregnancy, gestational week at measurement was coded as 0. When age at measurement was missing (12.1% for reticulocyte count, 19.1% for platelet count, 22.2% for HbA1c), data was imputed with multivariate imputation by chain equations including covariates. Ten complete imputed sets were analyzed and estimates were combined with `pool()`(R package ‘mice’ v3.13.0) (van Buuren and Groothuis-Oudshoorn, 2011). One-sided *p*-value were calculated as 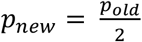 in case of directional agreement with the effect observed in the UKBB.

### 7. CNV burden analyses in the UKBB

#### CNV burden calculation

An individual’s CNV burden was defined as the number of Mb or genes affected by high-confidence autosomal CNVs (|QS| ≥ 0.5). For the later, CNVs overlapping exons, splice sites, non-coding RNA, 3’UTR, and 5’UTR (see *CNV region definition and annotation*) were retained and per sample lists of disrupted genes were compiled. Duplication and deletion burdens were calculated similarly, resulting in 6 burden metrices. Correlation between metrices was assessed with Pearson’s coefficient of correlation. Two-sided unpaired Wilcoxon rank sum test was used to assess differences in CNV burden between males and females.

#### CNV burden analysis

Linear regressions were performed between burden metrices and the 57 normalized covariate-corrected traits analyzed by CNV-GWAS. For sex-specific traits, samples from the opposite sex were excluded. Significance threshold was set at *p* ≤ 0.05/63 = 7.9×10^−4^, to account for the 6 life history traits tested (see *Life history*). To increase power, *mother’s* and *father’s age at death* were combined to assess the burden’ effect on parental lifespan. Linear regressions were computed between non-normalized but covariate-corrected traits and the burden to obtain effects on the unit scale. Results were meta-analyzed with GWAMA.

#### Burden analysis correction for modifier CNVRs

When assessing the impact of the CNV burden on a trait, CNVR associating with that trait under the mirror model were selected and a *sample × CNVR* matrix *G*, with value −1 (CNVR-overlapping (≥ 1 bp) deletion), 1 (CNVR-overlapping (≥ 1 bp) duplication), and 0 was generated. Effect of associated CNVRs was regressed out of the phenotype and burdens were corrected by subtracting the number of Mb/genes affected by the CNVR-overlapping CNV, before performing associations anew. To assess the impact of the duplication burden, CNVR found through the duplication-only model were considered and CNVR-overlapping deletions were set to 0 in *G*. To assess the impact of the deletion burden, CNVR found through the deletion-only model were considered and CNVR-overlapping duplications were set to 0 in *G*.

#### Fraction of inherited CNVs

To estimate the rate of CNVs inheritance, the fraction of shared CNVs among siblings was examined. Sibling pairs were identified as having a kinship coefficient between 0.2-0.3 and a fraction of SNPs with identity by state at 0 (IBS0) ≥ 0.0012 (Bycroft et al., 2018). We retained 16’179 pairs with one individual being among samples selected for the main CNV-GWAS (see *Sample selection*). Considering high confidence CNVs, shared CNVs were defined as either duplications or deletions on the same chromosome with ≥ 25 kb overlap. For each sibling pair, the fraction of CNVs the individual in the main analysis shared with his/her sibling was calculated (number of shared CNVs/total number of CNVs in that individual) and results were averaged over all pairs to obtain the mean fraction of shared CNVs. As a control, the analysis was repeated by pairing the 16’179 individuals from the main analysis with random individuals sampled without replacement from the remaining pool of individuals selected in *Sample selection*.

## Data Availability

Supplementary Figures and Tables are made available. Contact corresponding authors for access to the CNV-GWAS summary statistics.

## LIST OF ABBREVIATIONS

ALT: alanine aminotransferase
BAF: B allele frequency
BMI: body mass index
CMT: Charcot-Marie Tooth
CN: copy number
CNV: copy-number variant
CNVR: CNV region
CRP: C-reactive protein
EA: educational attainment
EstBB: Estonian Biobank
eQTL: expression quantitative locus
GGT: *γ*-glutamyl transferase
GW: genome-wide
GWAS: genome-wide association study
HbA1c: glycated hemoglobin
IBS0: identity by state at 0
ICD-10: International Classification of Diseases, 10^th^ Revision
LCR: low copy repeat
LDL: low-density lipoprotein
LRR: Log R ratio
MODY: maturity-onset diabetes of the young
TWMR: transcriptome-wide Mendelian randomization
PFB: population frequency of B allele
QC: quality control
QS: quality score
RCAD: renal cysts and diabetes
Rh: Rhesus
SCr: serum creatinine
SNP: single nucleotide polymorphism
UKBB: UK Biobank
WB: whole blood
WHR: waist-to-hip ratio

## ACKNOWLEDGMENTS

We thank all biobank participants for sharing their genetic and phenotypic data. UKBB computations were carried out on the JURA server, University of Lausanne. EstBB computations were performed in the High Performance Computing Center, University of Tartu. This work was supported by funding from the Department of Computational Biology (Z.K.) and the Center for Integrative Genomics (A.R.) from the University of Lausanne, as well as grants from the Swiss National Science Foundation (31003A_182632 to A.R.), Horizon2020 Twinning projects (ePerMed 692145 to A.R.), and the Estonian Research Council (PRG687, M.L. and R.M.). Critical reading of the draft by Johan Auwerx and Matthew Robinson was appreciated.

## AUTHOR CONTRIBUTIONS

C.A., A.R. and Z.K. conceived and designed the study; C.A. performed the main analyses, including the CNV calling, CNV-GWAS, burden analyses, and follow-ups on specific associations; M.L. and E.P. provided guidance for the CNV calling; E.P. provided guidance for association analyses; Z.K. supervised all statistical analyses; M.L. performed and R.M. supervised the replication study in the EstBB; M.C.S. performed the TWMR analyses under Z.K. and E.P.’s supervision; D.B. and M.S. collected and provided access to the CHUV maternity cohort data; M.P. performed analyses in the CHUV maternity cohort; C.A. drafted and A.R. and Z.K. made critical revisions to the manuscript; All authors read, approved, and provided feedback on the final manuscript.

## DATA AVAILABILITY

CNV-GWAS summary statistics in the UKBB are available as Supplementary data.

## WEB RESOURCES

- DatabaE of genomiC varIation and Phenotype in Humans using Ensembl Resources (DECIPHER) CNV Syndromes list, https://www.deciphergenomics.org/disorders/syndromes/list
- Genome Aggregation database (gnomAD) (v2.1.1), https://gnomad.broadinstitute.org/
- Genotype-tissue Expression (GTEx) project (v8) portal, https://gtexportal.org/home/
- Neale Lab UKBB genetic correlation, https://ukbb-rg.hail.is/
- Neale Lab UKBB summary statistics, http://www.nealelab.is/uk-biobank/
- Online Mendelian Inheritance in Man (OMIM), https://www.omim.org/
- ScyMed MediCalc, http://www.scymed.com/en/smnxfd/smnxfdad.htm

