## Supplementary Figures for "The individual and global impact of copy number variants on complex human traits"

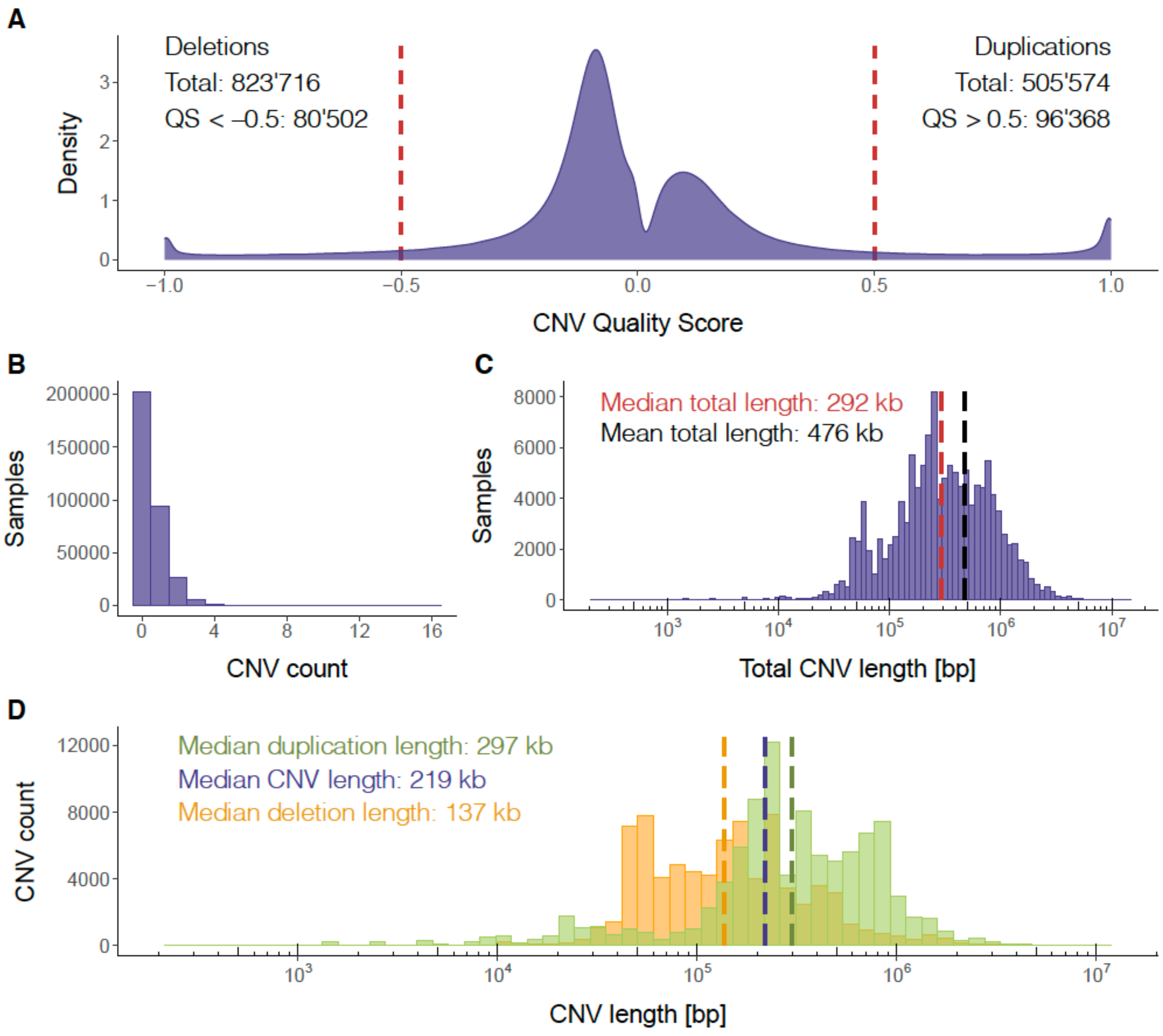


**Supplementary Figure 1 | Distribution of high confidence CNVs in the UKBB**

**(A)** Density plot of QS for the 1'329'290 called CNVs. High confidence duplications (QS ≥ 0.5) and deletions (QS ≤ -0.5), as indicated per the red dashed lines, were retained for downstream analyses. **(B)** Distribution of high confidence CNV counts per individual. **(C)** Distribution of the total amount of bases affected by high confidence CNVs per individual (logarithm scale); Red and black dashed lines show the median and mean total number of bases affected by CNVs among individuals with at least 1 CNV, respectively. **(D)** Distribution of high confidence duplications (green) and deletions (orange) length (logarithm scale). Dashed lines show the median duplication (green; 445kb), CNV (purple; 348 kb), and deletion (orange; 232 kb) length.

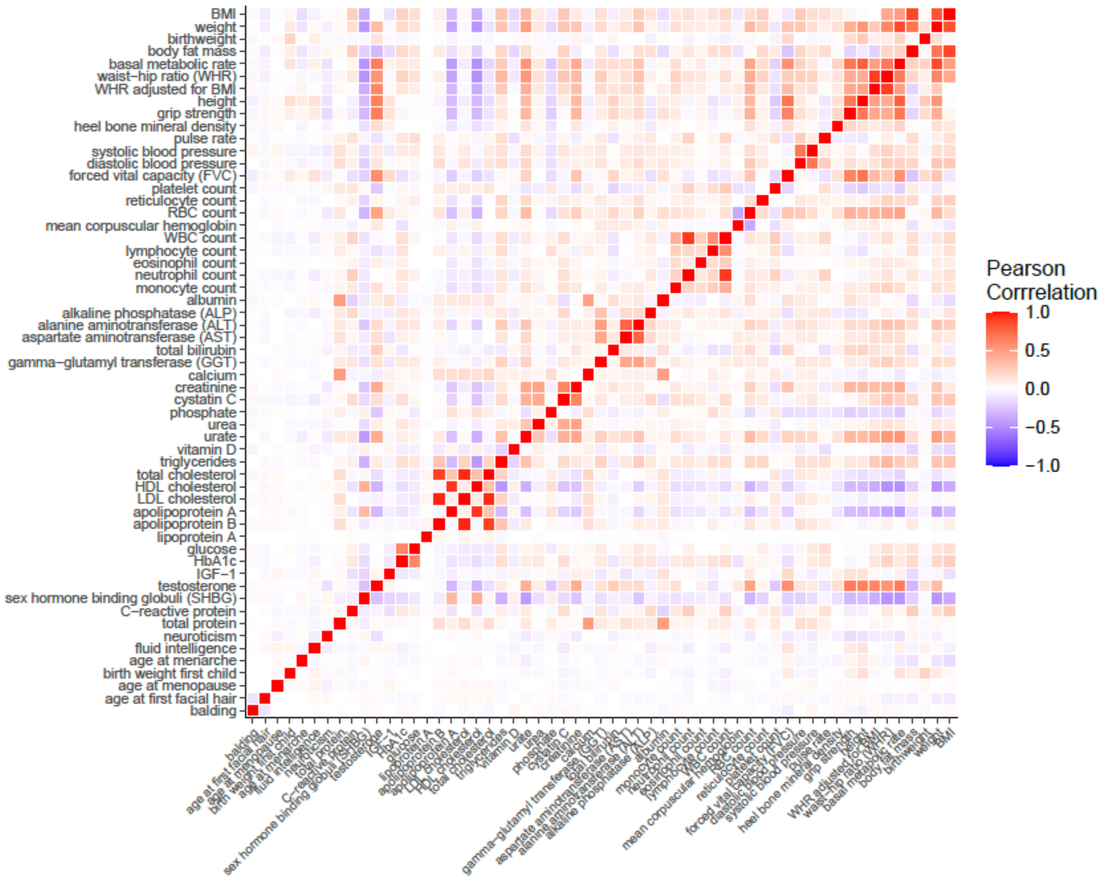


**Supplementary Figure 2 | Assessed complex traits**

Pearson correlation across the 57 continuous traits assessed by CNV-GWAS.

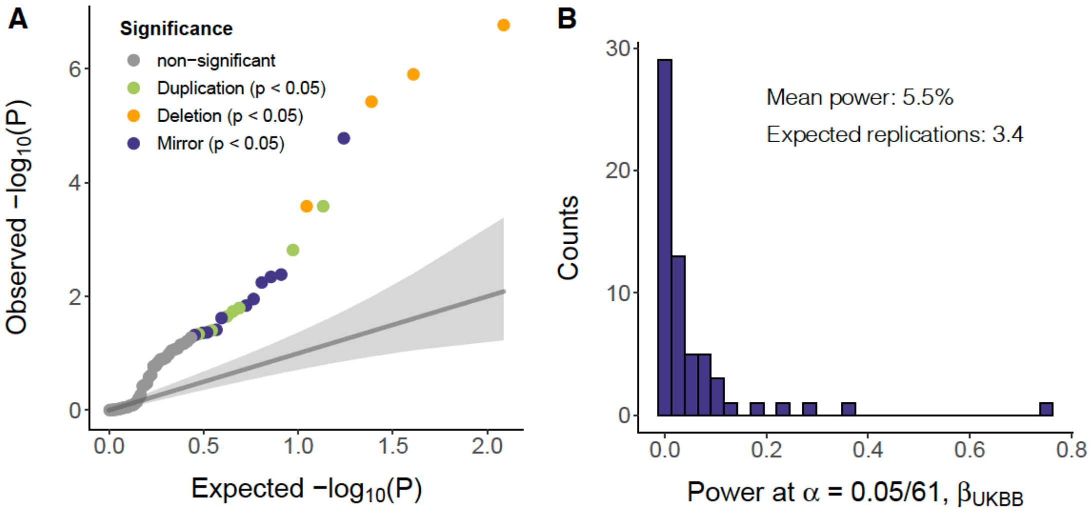


**Supplementary Figure 3 | Replication of CNV-GWAS signals in the EstBB**

**(A)** Expected versus observed negative logarithm of *p*-values for the 61 CNV-trait pairs assessed in the EstBB. Directionally concordant and nominally significant ($p \leq 0.05$) signals replicated with the duplication-only, deletion-only, or mirror association model are in green, orange, or purple, respectively. Non-significant signals ($p > 0.05$) are in grey. Shade represents the 95% confidence interval around expected values. **(B)** Distribution of replication power at $\alpha = 0.05/61 = 8.2 \times 10^{-4}$, assuming similar effect sizes to the one observed in the UKBB, for the 61 signals replicated in the EstBB. The mean power is 5.5%, corresponding to an expected number of multiple-testing survival replications of 0.055*61 = 3.4.

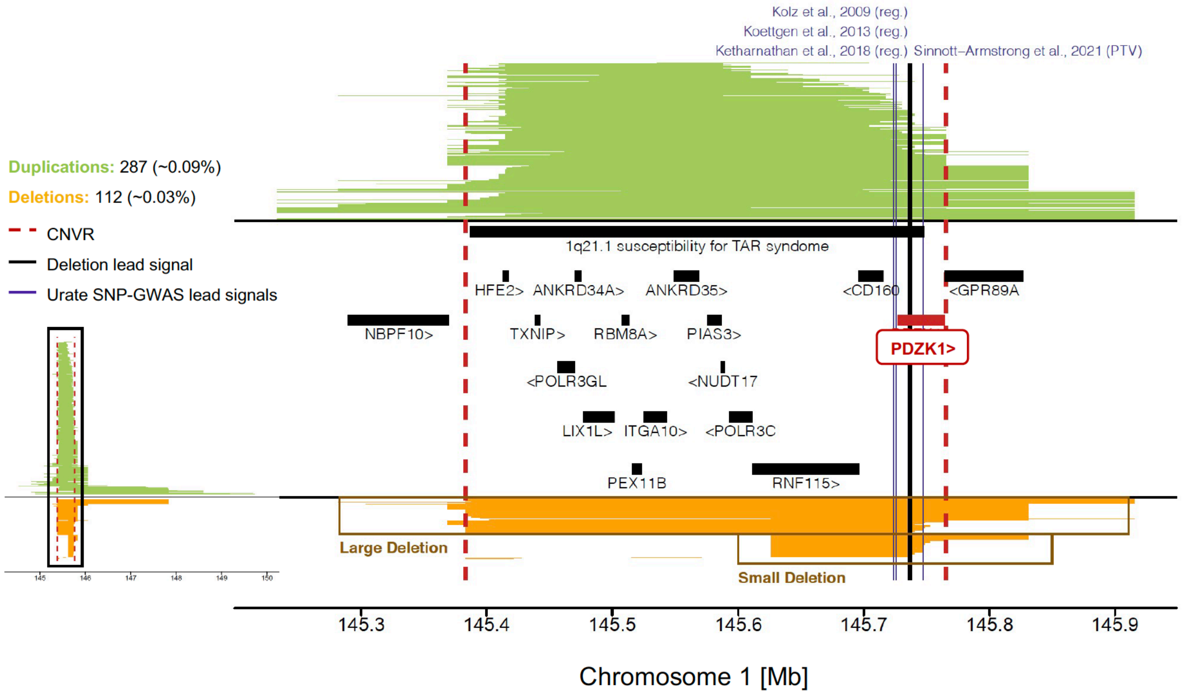


### Supplementary Figure 4 | 1q21.1 deletion and decreased serum urate

Mapping of CNVs overlapping the 1q21.1 deletion region associated with decreased serum urate. Number and frequency of duplications and deletions are at the top left; The left plot shows all overlapping CNVs and the right plot focuses on the central CNVR represented by red dashed lines (chr1:145'383'239-145'765'206). Duplications are in green, deletions in orange; Black line indicates the lead signal for serum urate (deletion-only); Purple lines indicate serum urate-associated SNPs (reg. = regulatory variant; PTV = protein-truncating variant). Recurrent CNV and overlapping protein-coding genes are shown in black, with the exception of *PDZK1* in red. Boxes separate large vs. small deletion carriers.

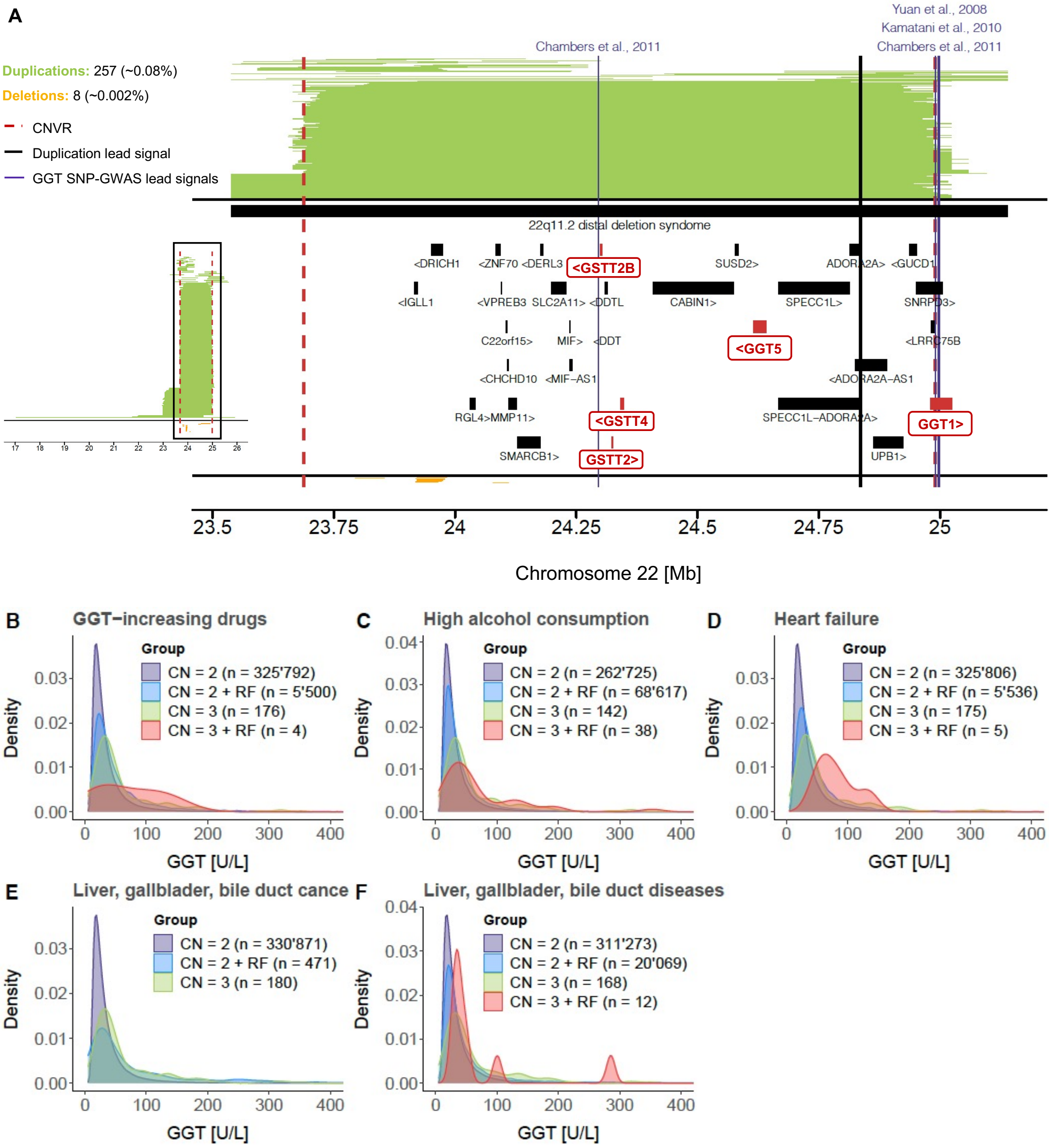


**Supplementary Figure 5 | 22q11.23 duplication and increased serum GGT**

**(A)** Mapping of CNVs overlapping the 22q11.23 duplication associated with increased GGT. Number and frequency of duplications and deletions are at the top left; The left plot shows all overlapping CNVs and the right plot focuses on the central CNVR represented by red dashed lines (chr22:23'688'345-24'990'213). Duplications are in green, deletions in orange; Black line indicates the lead signal for GGT (duplication-only); Purple lines indicate GGT-associated SNPs. Recurrent CNV (truncated) and overlapping protein-coding genes are shown in black, with the exception of genes involved in glutathione metabolism in red. **(B-F)** Density plot showing the distribution of GGT levels in copy-neutral (CN = 2) and duplication carriers (CN = 3) with or without various risk factors (RF) for increased GGT: **(B)** GGT-increasing drugs, **(C)** high alcohol consumption, **(D)** heart failure, and **(E)** cancer or **(F)** other diseases of the liver, gallbladder, or bile ducts. The sample size for each category is indicated. Plots were truncated to 400 U/L (maximum value: 1167 U/L).

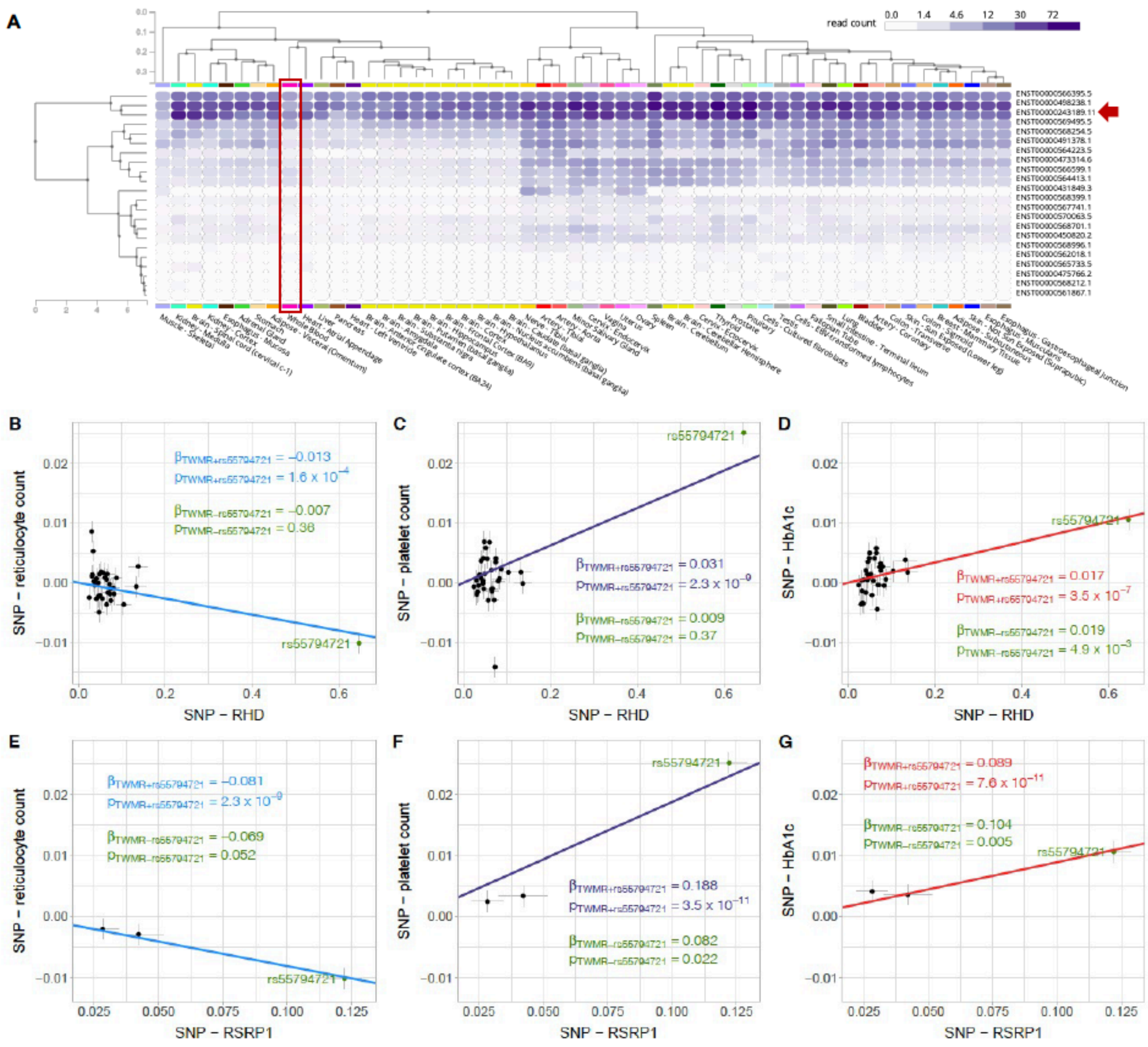

**Supplementary Figure 6 | 1p36.11 deletion and altered hematological traits**

**(A)** GTEx v8 isoform expression for *RSRP1* in 54 tissues. WB is circled in red, with the arrow pointing at the isoform with the highest expression in WB, ENST00000243189. **(B-G)** SNP-exposure (x-axis) versus SNP-outcome (y-axis) plots for TWMR analyses estimating the causal effect of **(B)** *RHD* → reticulocyte count, **(C)** *RHD* → platelet count, **(D)** *RHD* → HbA1c, **(E)** *RSRP1* → reticulocyte count, **(F)** *RSRP1* → platelet count, and **(G)** *RSRP1* → HbA1c. Grey horizontal and vertical lines represent the standard error around the SNP-exposure and SNP-outcome estimates, respectively. Causal effect estimates and associated *p*-values are reported for each analysis. The outlier SNP rs55794721 is in green. Causal effects were re-estimated omitting rs55794721 and are reported in green.

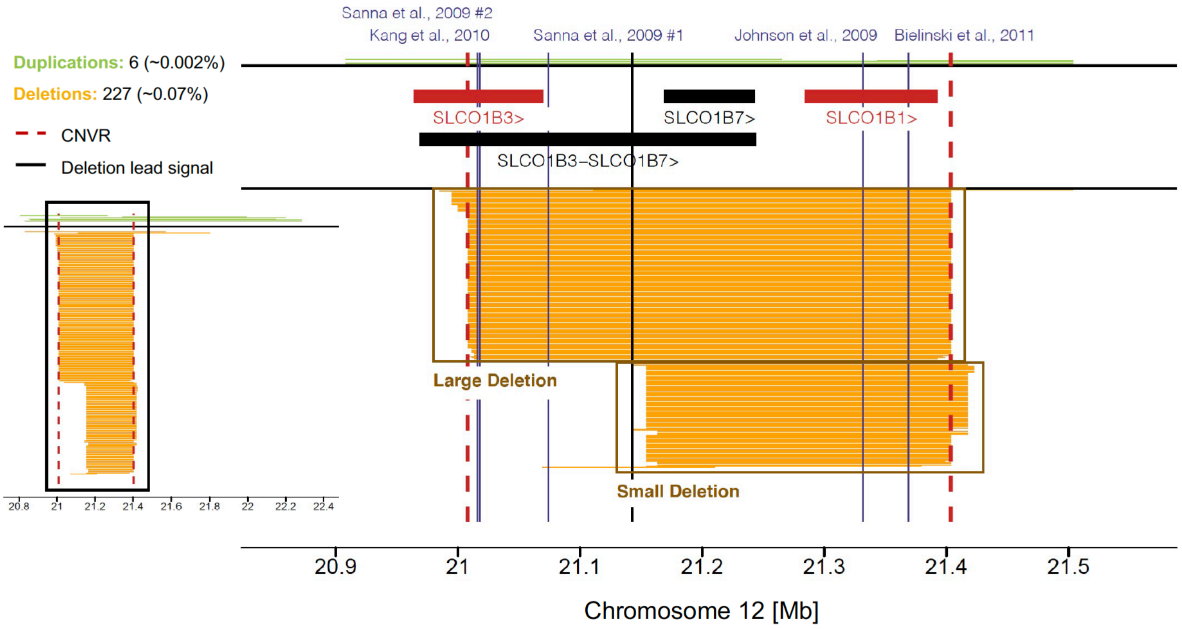


**Supplementary Figure 7 | 12p12.2-p12.1 deletion and increased total bilirubin**

Mapping of CNVs overlapping the 12p12.2-p12.1 deletion associated with increased total bilirubin. Number and frequency of duplications and deletions are at the top left; The left plot shows all overlapping CNVs, the right plot focuses on the central CNVR represented by red dashed lines (chr12:21'008'080-21'403'457). Duplications are in green, deletions in orange; Black line indicates the lead signal for total bilirubin (deletion-only); Purple lines indicate serum bilirubin-associated SNPs; Overlapping protein-coding genes are shown in black, with the exception of Rotor syndrome-associated genes *SLCO1B1* and *SLCO1B3* shown in red. Boxes separate large vs. small deletion carriers.

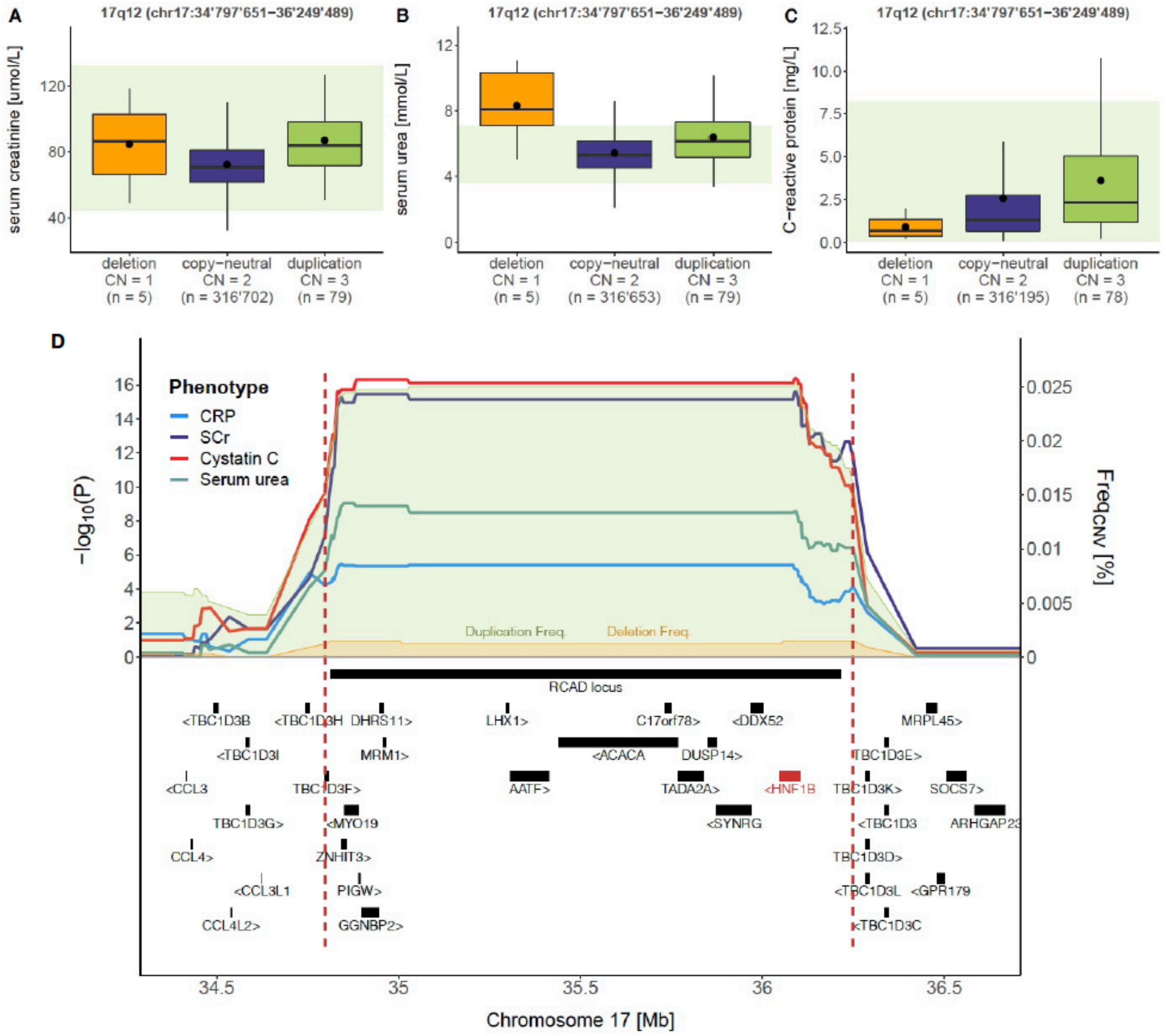


**Supplementary Figure 8 | 17q12 CNVs and renal phenotypes**

Boxplots representing levels of **(A)** SCr, **(B)** serum urea, and **(C)** CRP in individuals with 17q12 deletion, copy-neutrality, or duplication. CNVR coordinates are at the top; Copy number (CN) and sample size are reported for each category; Dots show the mean; Outliers are not shown; Green bands show normal clinical range for **(A)** SCr: 44.2-132.6 µmol/L, **(B)** serum urea: 3.6-7.1 mg/L, and **(C)** CRP: 0.07-8.2 mg/L. **(D)** Association plot for the 17q12 CNVR. Red dashed lines delimit the duplication-only associated CNVR (chr17:34’797’651-36’249’489); Left y-axis shows the negative logarithm of association *p*-value for CRP (blue), SCr (purple), cystatin C (red), and serum urea (turquoise); Right y-axis shows CNV frequency [%], with duplication frequency in green and deletion frequency in orange; Overlapping recurrent CNVR and genes are shown in black, except for the putative causal gene for the observed associations, *HNF1B*, shown in red.

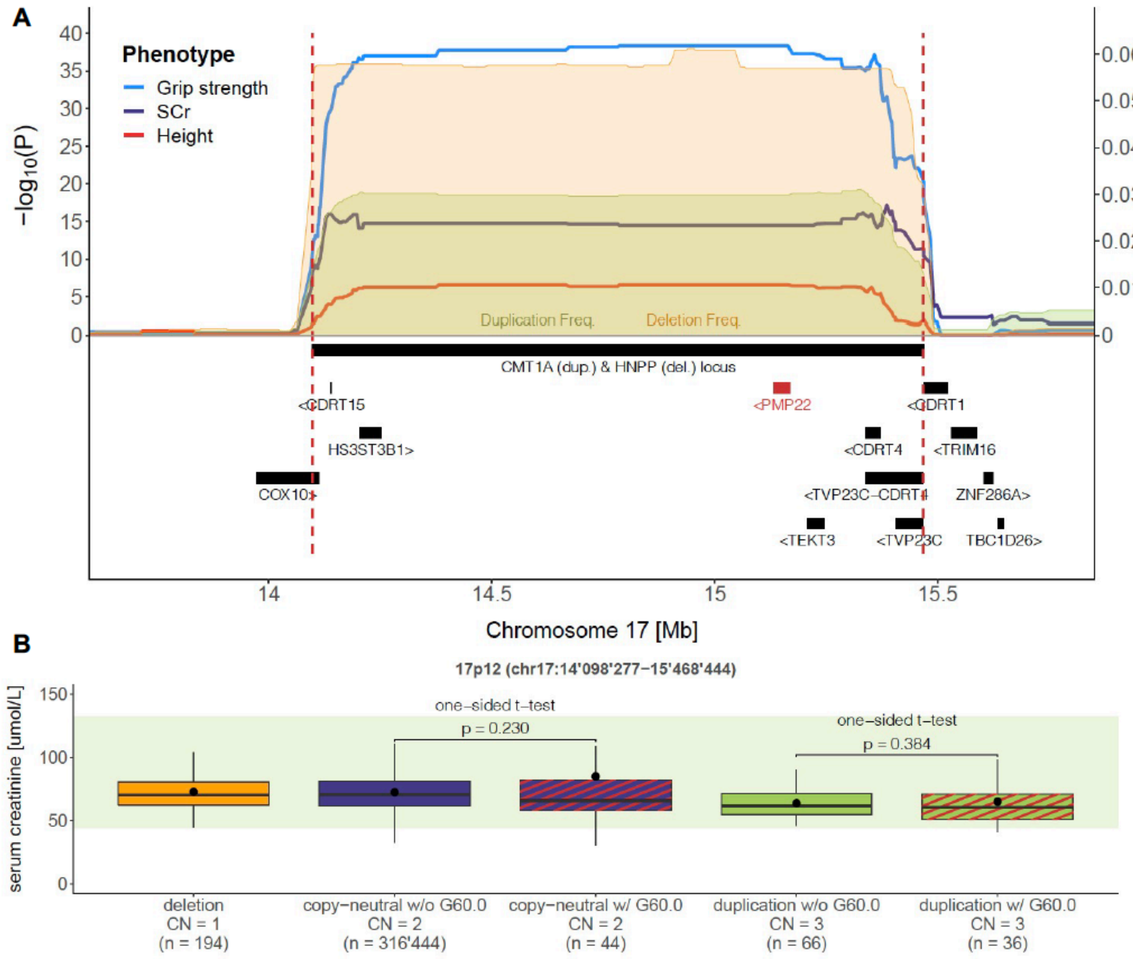


**Supplementary Figure 9 | 17p12 duplication and muscle phenotypes**

**A)** Association plot for the 17p12 CNVR. Red dashed lines delimit the duplication-only associated CNVR (chr17:14’098’277-15’468’444); Left y-axis represents the negative logarithm of association *p*-value for hand grip strength (blue), SCr (purple), and height C (red); Right y-axis represents CNV frequency [%], with duplication frequency in green and deletion frequency in orange; Overlapping recurrent CNVR and genes are shown in black, except for the putative causal gene for the observed associations, *PMP22*, shown in red. **(B)** Boxplots representing SCr levels of individuals with 17p12 deletion, copy-neutrality, or duplication, the two latter being split according to the presence (w/) or absence (w/o) of a hereditary motor or sensory neuropathy diagnosis (ICD-10 G60.0; Red stripes). CNVR coordinates are at the top; Copy number (CN) and sample size are reported for each category; Dots show the mean; Outliers are not shown; Green bands show normal clinical range for SCr: 44.2-132.6 µmol/L.
